# NOX4 contributes to the initiation and progression of AAA in a cell type-specific manner

**DOI:** 10.64898/2026.03.04.26347161

**Authors:** Anja Hofmann, Anupam Sinha, Christoph Schürmann, Bianca Hamann, Maria Sabater-Lleal, Franziska Horn, Marvin Kapalla, Margarete Müglich, Irakli Kopaliani, David M. Poitz, Albert Busch, Ralph A. Bundschuh, Henning Morawietz, Christian Reeps, Katrin Schröder

## Abstract

**Background:** Abdominal aortic aneurysm (AAA) is a disease with altered vessel wall architecture and integrity. AAA rupture is associated with high mortality. Reactive oxygen species, such as those produced by members of the NADPH oxidase (NOX) family, play a central role in several aspects of vascular physiology. In particular, the role of NOX4 appears to be highly cell and context specific.

**Methods:** This study analyzed the role of NOX4 in late-stage human AAA specimen and in Nox4−/− mice with experimentally induced AAA.

**Results:** NOX4 expression was reduced in human AAA. In a mouse model of AAA, loss of Nox4 conferred protection against AAA formation, suggesting a pathogenic role. Single cell analysis of human AAA revealed that NOX4 is primarily expressed in fibroblasts, s.mooth muscle, and endothelial cells. NOX4 mRNA expression was strongly associated with ECM synthesis and ECM remodeling pathways. Angiogenic signatures were reduced in AAA, and sub-cluster analysis of endothelial cells identified two major groups: microvascular and lymphatic endothelial cells (LEC), with very low NOX4 expression in LEC. Quantification of the *vasa vasorum* revealed a shift in vessel size distribution, with a reduction in the number of small vessels (<8 µm) and an increase in large vessels (>26 µm) correlating with increasing aortic diameter. Markers of lymphangiogenesis, including VEGFC and PROX1, were upregulated in AAA. Pseudotime trajectory analysis suggested transdifferentiation of LECs into myofibroblasts, a process associated with increased NOX4 mRNA expression.

**Conclusion:** NOX4 plays a role in the pathogenesis of AAA and is primarily expressed in fibroblasts, smooth muscle cells, and endothelial cells. Single-cell and pseudotime analyses revealed that NOX4 is associated with ECM remodeling, reduced angiogenic signatures, and the transdifferentiation of lymphatic endothelial cells into myofibroblasts.

**Clinical Perspective:** *What is new?:*

- In human AAA, NOX4 is associated with pro-fibrotic effects.
- NOX4 appears to play a central role in cell differentiation processes in AAA, supporting the expansion of the fibroblast population.
- The percentage of small microvessels (<8 µm) is increased in human AAA, and NOX4 expression correlates positively with the proportion of small vessels.
- The cell-cell communication network of endothelial cells in AAA appears to have a profile that supports fibrosis.
- Lymphatic endothelial cells and markers of lymphangiogenesis were found in AAA.
- Lymphatic endothelial cells transdifferentiate into myofibroblasts, a process accompanied by increased NOX4 expression.
- NOX4 may serve as a mechanistic link between lymphangiogenesis and fibrosis, bridging vascular remodeling and fibrotic progression.

*Translational Perspective?:*

- Targeting NOX4 represents a promising therapeutic strategy for mitigating fibrotic remodeling in late-stage AAA.
- Targeting the specific receptors mediating the interaction between lymphatic endothelial cells, fibroblasts, and inflammatory cells may reveal novel therapeutic targets.

**Graphical abstract:** 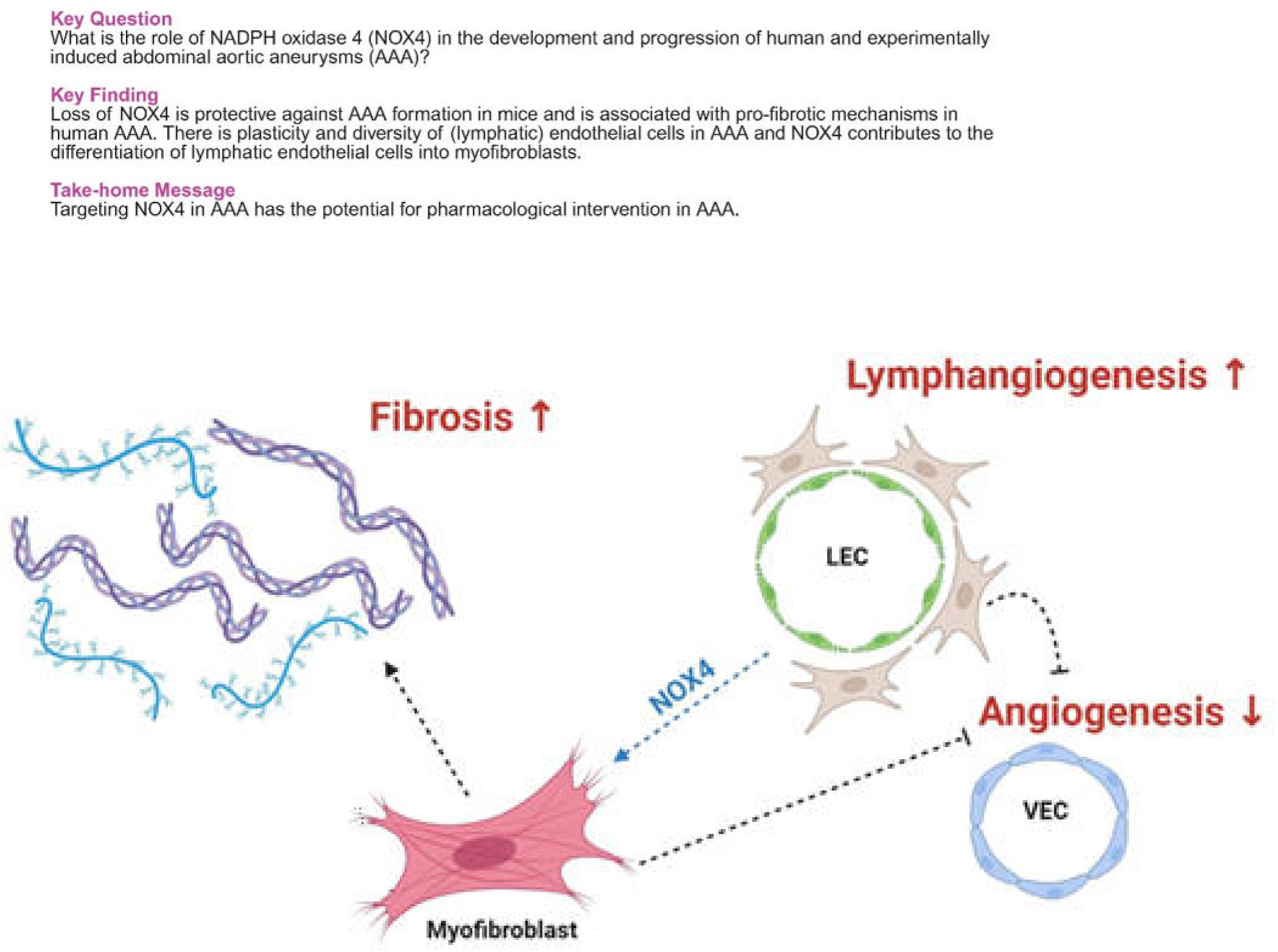

## Introduction

Abdominal aortic aneurysm (AAA) is a common vascular disease defined as a dilation of the abdominal aorta >30 mm affecting mostly men >65 years.^1^ Over time, the diameter and the risk of rupture of the AAA increases. Destruction and remodeling of components of the vessel wall are thought to be a main reason for AAA development and rupture. The underlying mechanisms are not well-understood. Accordingly, conservative risk factor management and surgical interventions are the only treatment options so far.^2^ A main characteristic of AAA is the pathological remodeling of the aortic extracellular matrix (ECM). Although the total collagen content is high in AAA, an impaired collagen homeostasis and defects in collagen microarchitecture may promote nonfunctional remodeling.^3^ Other components of the ECM such as proteoglycans, growth factors and proteases, may also play a role. They may be released as a secondary response to an initial injury to stimulate vascular repair processes. Upon activation, fibroblasts transform into myofibroblasts, which express alpha-smooth muscle actin. They migrate and proliferate, produce ECM and secrete pro-inflammatory cytokines. Eventually, they replace or substitute for vascular smooth muscle cells, which disappear during AAA formation.^4^ Myofibroblasts initiate wound healing and facilitate the formation of a fibrotic scar.^5^ Importantly, myofibroblasts can transdifferentiate not only from fibroblasts but also from endothelial cells.^5^ Transdifferentiation is induced by cytokines, chemokines and growth factors in conjunction with increased wall tension,^5^ suggesting that cells other than myofibroblasts may be involved. In fact, the tunica adventitia contains immune cells, lymphatic vessels, and endothelial cells in the *vasa vasorum*.^4^ NADPH oxidase 4 (NOX4) has been shown to contribute to the differentiation of adipocytes, osteoblasts and endothelial cells and to switch the phenotype of smooth muscle cells and macrophages.^6^ ^7^ Eventually, NOX4 mediates the transdifferentiation of murine adventitial fibroblasts into myofibroblasts.^8^ In adventitial fibroblasts, NOX4 is expressed at high levels and produces H_2_O_2_, which may induce cytokine secretion.^9^ SMC-specific deletion of Nox4 has been shown to reduce aortic collagen and MMP2 expression.^10^ Additionally, systemic Nox4 deletion nearly abolished angiotensin II-induced AAA formation in hph1/Nox4 double knockout mice.^11^ However, in contrast to its protective role in AAA, Nox4 deletion promotes atherosclerosis and is generally considered detrimental in the cardiovascular system.^12,13^ Despite the existence of tissue- and cell-specific Nox4 knockout mice, the exact cells that express NOX4 in AAA have not yet been identified. Identifying cell-specific, disease-associated transcriptional programs may offer critical insights and novel opportunities for the development of targeted therapies for AAA. However, translating findings from animal models to human disease remains challenging. In this study, we investigated the role of NOX4 in AAA formation and progression by analyzing tissue from patients with end-stage AAA and employing a complementary murine model of the disease.

## Materials and Methods

The authors declare that all supporting data are available within the article and its online supplementary files. All Materials and Methods are described in the supplementary files.

Male mice aged 8–10 weeks were used to induce abdominal aortic aneurysms (AAAs) via a combination of systemic Angiotensin II (AngII) infusion and dietary modulation. AngII was delivered using Alzet osmotic minipumps (model 2001, Cupertino, USA), which were implanted subcutaneously. Prior to pump implantation, mice were anesthetized via nasal administration of 2–3% isoflurane in oxygen.

## Results

### Aortic NOX4 mRNA expression is lower in AAA and associated with changes in the ECM

NOX4 mRNA expression was analyzed in apparantly healthy vessel segments (n=6), electively treated (n=46) and ruptured human AAA (n=19). NOX4 was decreased in eAAA and tended to be increased in rAAA (**Fig. 1A**), consistent with published studies^14^. Supporting a detrimental role for increased NOX4 in ruptured AAA, NOX4 mRNA expression was positively correlated with cleaved caspase-3 positive areas, elastin degradation and DNA binding activity of NF-κB (**Supplementary Table 1**). NOX4 mRNA expression decreased with increasing AAA diameter (rp=-0.469, *P*=.006) (**Fig. 1B**). Furthermore, the relationship between NOX4 mRNA expression and histopathological features of the AAA was analyzed using NOX4 as a response variable in relation to changes in aortic wall composition. NOX4 mRNA expression was reduced in samples with high CD31 expression, suggesting anti-angiogenic mechanisms (**Fig. 1C**). Fibrillar collagens type I and type III account for 80-90 % of all collagens in the aorta.^15^ NOX4 mRNA expression was increased in AAA specimens with high COL1A1 and COL3A1 expression, supporting a pro-fibrotic role (**Fig. 1D,E**). Loss of NOX4 is associated with an increased aortic inflammation^12^ and in AAA high mRNA expression of the pro-inflammatory cytokine IL6 was associated with increased NOX4 mRNA expression (**Fig. 1F**).

**Figure 1:**
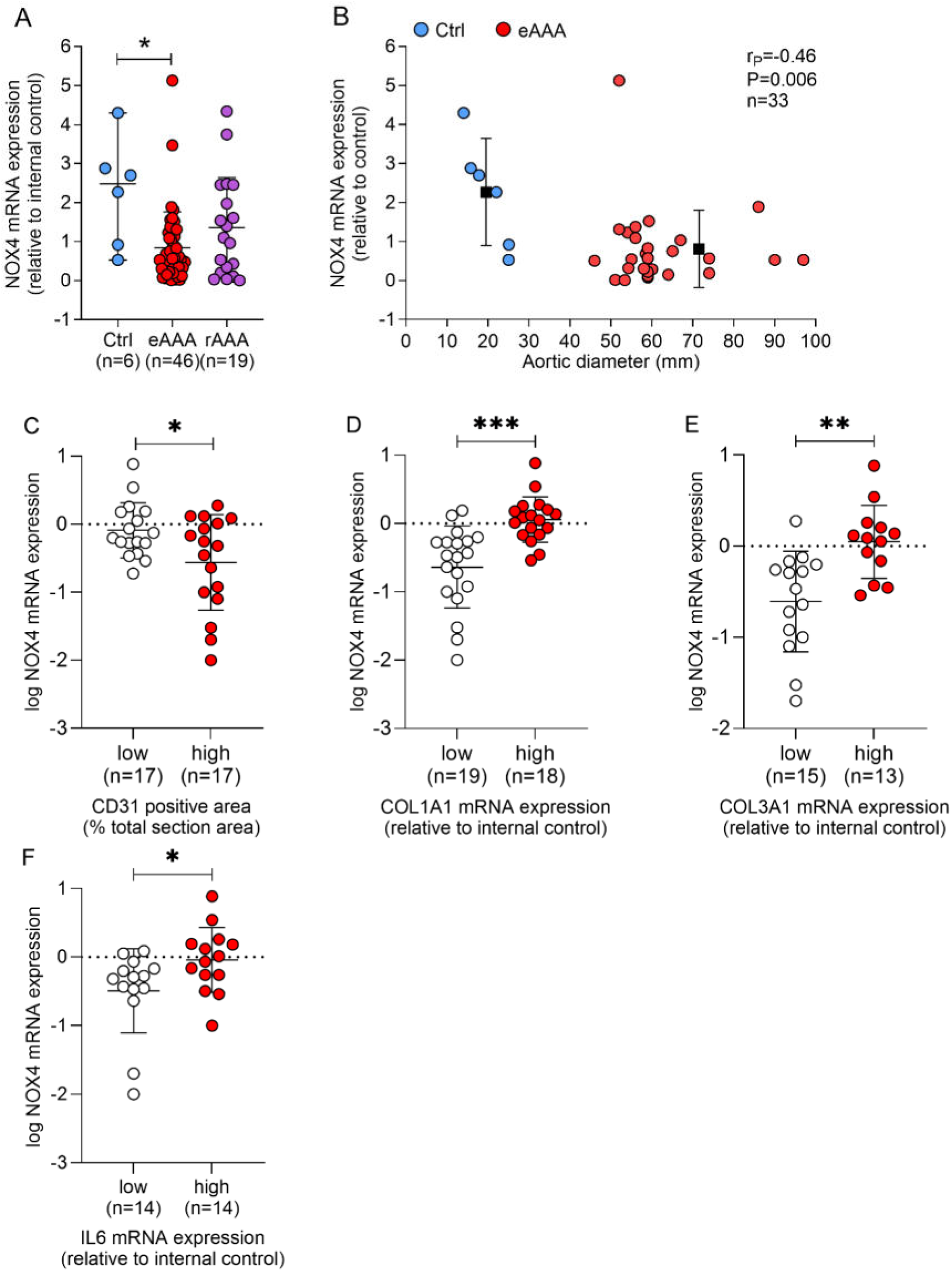
Aortic NOX4 mRNA expression, and associations with AAA diameter, collagen expression, endothelialisation and inflammation in electively treated AAA. Aortic specimens were obtained from patients undergoing elective (eAAA, red dots) surgical repair, patients with ruptured AAA (rAAA, violet dots) and non-diseased controls (Ctrl, blue dots). **A,** Comparison of NOX4 mRNA expression in Ctrl, eAAA and rAAA. NOX4 expression was analyzed by qPCR and data are normalized to an internal control (=1). **B,** Correlation of NOX4 mRNA expression with AAA diameter in aortic segements obtained from non-diseased controls (blue dots) and in electively treated AAA (red dots). **C,** CD31 protein expression was visualized by immunohistochemistry and quantified. **D,** COL1A1, **E,** COL3A1 and **F,** IL6 mRNA expression was analyzed by qPCR. Data are normalized to an internal control (=1). The expression of all markers was divided into “low” or “high” by using the median of each data set. **Statistics:** Statistically significant outliers were detected by Grubb’s outlier test and one sample in the eAAA group (Fig. 1A) was excluded from the analysis. **A,** Kruskal-Wallis and Dunn’s *post hoc* test. \*\**P*<.001 Ctrl *vs.* eAAA. **B,** Pearson’s correlation rP. **C-F,** Due to scatter, NOX4 mRNA expression was logarithmically transformed and data are presented as a scatter dot plot with the horizontal line depicting the mean. Data were compared using the Student’s T-test. \**P*<.05, \*\**P*<.001, \*\*\**P*<.0001 low *vs* high. Controls (n=6), electively abdominal aortic aneurysm (eAAA; n=46), and ruptured AAA (rAAA; n=19) samples were included in the analysis. All samples represent biological replicates.

### Nox4 deletion protects from AAA formation in mice

To analyze whether the decrease in NOX4 mRNA expression with increasing AAA diameter is associated with protective or detrimental effects, we analyzed a mouse model with genetic deletion of NOX4. In a model of AngII+BAPN induced AAA formation deletion od Nox4 was protective (AAA in WT 80% *vs.* AAA in Nox4−/− 20%) and the maximal diameter of the AAA was smaller in Nox4−/− mice (**Fig. 2A-E**). Absolute adventitial collagen content was reduced in the aorta of Nox4−/− mice (**Supplementary Fig. 1A-C**). Collagen deposition as a sign of vascular fibrosis and was decreased in the absence of Nox4. In contrast, Nox4 knockout prevents elastin strand breaks while the media thickness was similar in wildtype and Nox4−/− mice (**Supplementary Fig. 1D-F**). The data support a detrimental role of Nox4 in aneurysm formation. However, mouse models are artificial and may not reflect the human situation. Therefore, we focused on human AAA specimens for further analysis.

**Figure 2:**
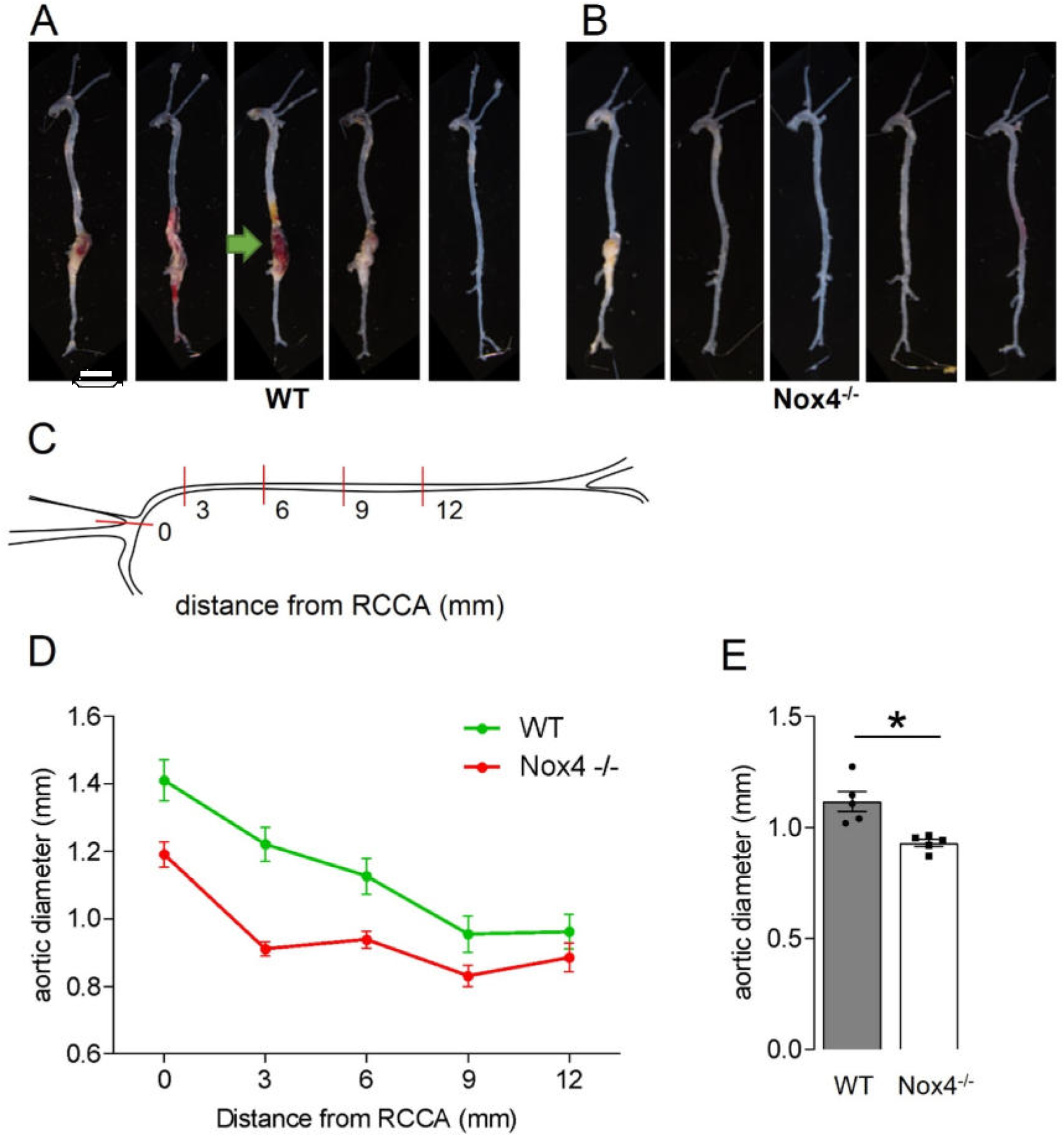
Nox4 deletion protects mice from AAA formation. WT and Nox4^-/-^ mice were treated with a combination of AngII (1 mg/kg/ day) and a high-fat diet for 4 weeks. AAA formation was additionally induced by BAPN administration in the drinking water (1 mg/mL) during the first 2 weeks of treatment. Representative macroscopic images of aortas from **A,** WT and **B,** Nox4−/− mice with arrows indicating the AAA. **C,** The figure schematically shows the measuring points along the aorta, starting at the right common carotid artery (RCCA). **D,** Maximal aortic diameter measured by ultrasound after 28 days of treatment at the indicated distances from the RCCA in WT (green) and Nox4−/− mice (red). **E,** Average of all aortic diameters. Statistics: Student’s T-test. Data are presented as mean±SD. \**P*<.05 WT *vs.* Nox4−/−. n=5 animals per group. Scale bar: 5 mm.

### NOX4 is linked to fibrotic and anti-angiogenic mechanisms

Single cell analysis of aortic segments from four patients suffering from infrarenal AAA (**Supplementary Fig. 2**) and unsupervised clustering resulted in nine distinct major cell partitions (**Supplementary Fig. 3A,B**). The distribution ratio of the different cell partitions were highly patient specific, validating puplished data of late-stage human AAAs.^16^ Plotting the top marker genes of each cluster unmasked several subclusters (**Supplementary Fig. 3C,D**). For all further analyses we focused on non-immune cells and used a published dataset as controls.^17^ NOX4 expression in AAA was highest in fibroblasts followed by smooth muscle and endothelial cells, while controls showed the highest expression in smooth muscle cells (**Supplementary Fig. 4**). We analyzed genes differentially expressed depending on the presence or absence of NOX4 (**Fig. 3A**). NOX4 expression was associated with high expression of genes involved in ECM organisation and fibrosis, supporting the data in Nox4−/− mice and the histopathology in human AAA. Further, NOX4 expression was associated with GO and KEGG terms involved in ECM organisation, migration and differentiation. In contrast, endothelial barrier function, tight junction and chemokine signaling were reduced in cells that express NOX4 (**Fig. 3B-E**). The gene expression pattern was used to estimate transcription factor activity in NOX4-expressing *vs.* non-NOX4-expressing cells. We identified clusters belonging to differentiation, pluripotency/stemness, fibrosis to be upregulated in the presence of NOX4. In contrast, transcription factors for TGF-β1 signaling inhibitors and pro-angiogenic genes were down regulated (**Supplementary Fig. 5B**).

**Figure 3:**
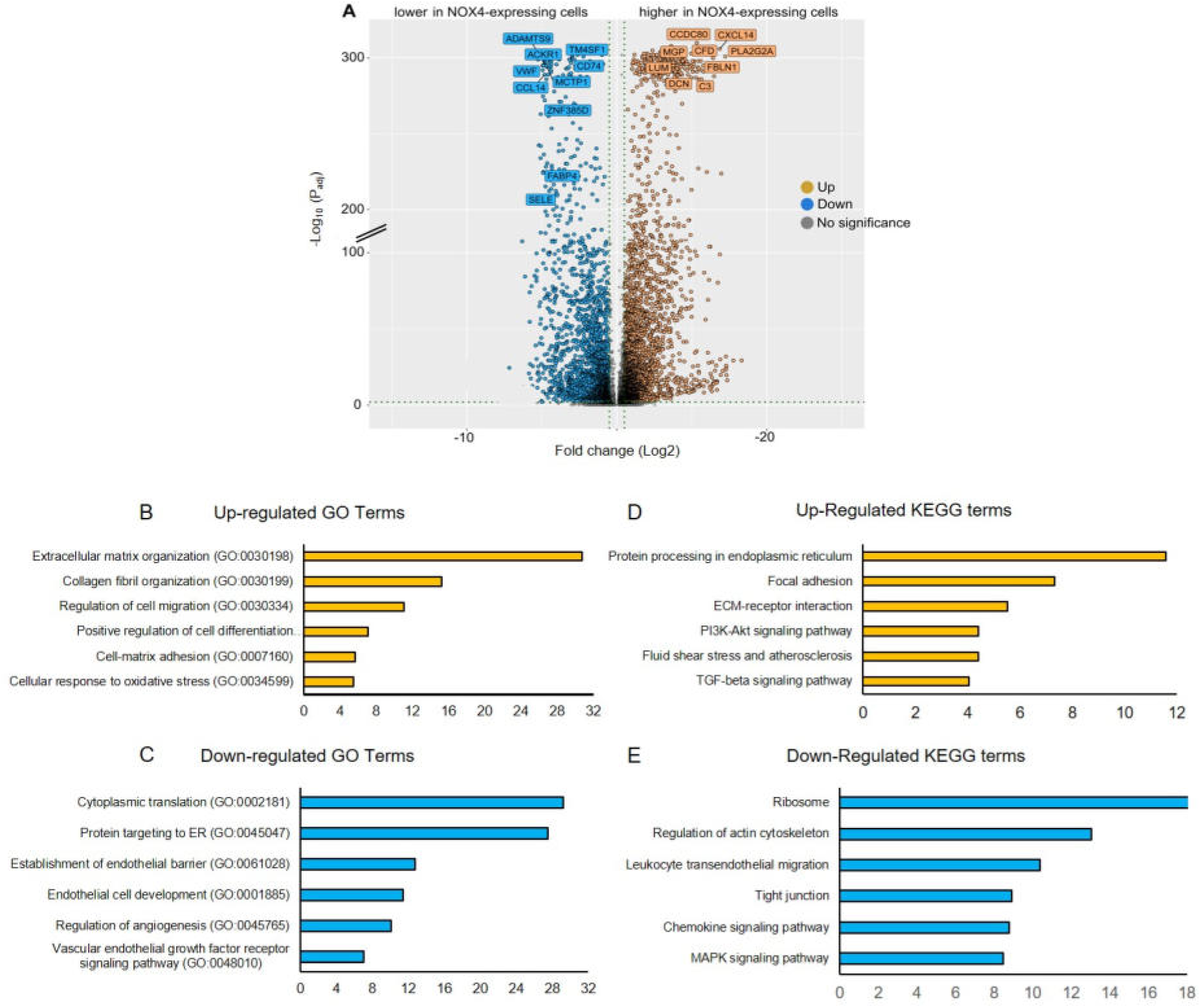
NOX4 expression and differential expression analysis and functional annotation in NOX4-expressing cells in human AAA. **A,** Volcano plot of the top differentially expressed genes in NOX4-expressing *vs.* non-NOX4-expressing cells within all non-immune cells. The left side shows negative regulation of genes and the right side shows the positive regulation of genes. The log2-fold differences between the two groups are plotted on the x-axis and the -log10 *P*-value differences are plotted on the y-axis. The dotted vertical lines represented the q-value 0.5 and <−0.5 corresponding to upregulated and downregulated genes, respectively. The dotted horizontal line is the threshold above which significant differential expressed genes are located. **B, C,** Gene ontology (GO) and **D, E,** Kyoto Encyclopedia of Genes and Genomes (KEGG) pathway enrichment analysis of differentially expressed genes in NOX4-expressing cells. The size of the dot plot is based on the gene count enriched in the pathway (fold difference *vs.* non-NOX4-expressing cells), and the dot color depicts the FDR-corrected log10 *P*-values. Biological processes are ranked and the most relevant are highlighted in red and less significant in blue. Electively treated abdominal aortic aneurysm (eAAA; n=4) were included in the analysis. All samples represent biological replicates.

In conclusion, the data suggest that matrix synthesis, along with pathways and transcription factors central to fibrosis, including those involved in matrix remodelling and stabilisation, are enriched in NOX4-expressing cells, whereas angiogenic mechanisms appear to be diminished.

### Smooth muscle cells and fibroblast (de)differentiation contributes to fibrotic remodeling in human AAA

Within the fibroblast cluster, we identified NOX4 in adventitial fibroblasts, myofibroblasts, progenitor, and a population of cells that may have differentiated from mesenchymal or haematopoietic cells, such as TGF-β1 activated fibrocytes (**Fig. 4A-D, Supplementary Fig. 6A**). GO and KEGG analyses verified pro-fibrotic alterations as well as myofibroblast differentiation in NOX4-expressimg subclusters (**Supplementary Tables 2-6**). Staining of AAA crosssections for fibroblast-activated protein (FAP) confirmed the presence of activated fibroblasts. Previous studies have shown colocalisation of FAP with α-SMA and suggested FAP as a marker of myofibroblast activation in AAA ^18^. Loss of contractile smooth muscle cell markers is a hallmark of end-stage AAA ^19^. Indeed, we identified smooth muscle cell subclusters that belong to myofibroblasts/fibroblast-like and “osteogenic-like smooth muscle cells”, both expressing NOX4 (**Fig. 4F-I, Supplementary Fig. 7A, Supplementary Table 6**).

**Figure 4:**
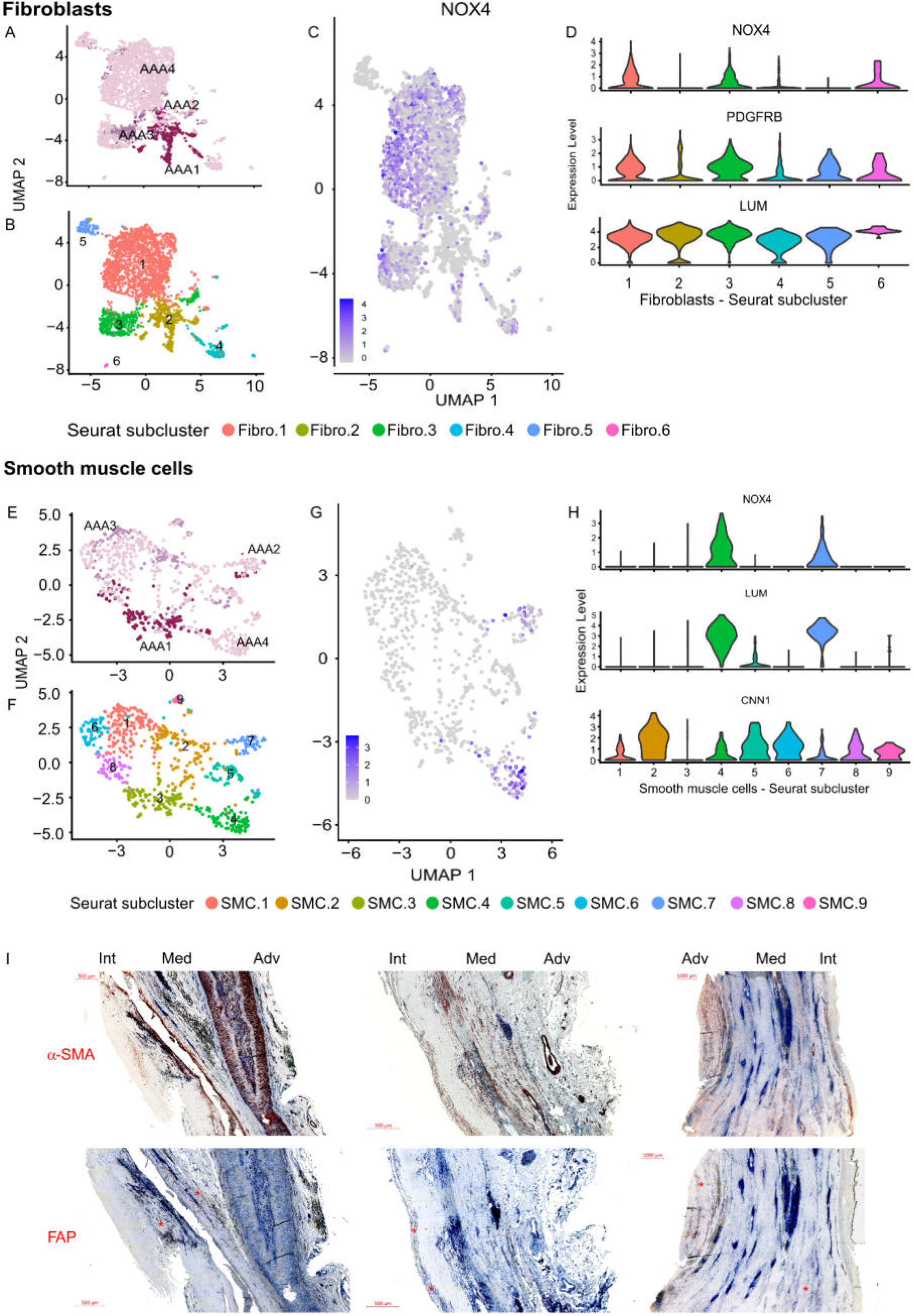
Characteristics of fibroblast and smooth muscle Seurat subclusters in human AAA. **Fibroblasts: A,** Uniform Manifold Approximation and Projection (UMAP) plot of cells from four different patients with AAA (AAA.1-AAA.4). The colors denote the different AAA samples. **B,** UMAP plot of the different cell clusters within each AAA. The colors and numbers denote the different cell types identified by subclustering of the major cell types. **C,** feature blots of NOX4 and **D,** violin plots for relative expression of NOX4, PDGFRB and LUM in each subcluster. **Smooth muscle cells: E,** Uniform Manifold Approximation and Projection (UMAP) plot of cells from four different patients with AAA (AAA.1-AAA.4). The colors denote the different AAA samples. **F,** UMAP plot of the different cell clusters within each AAA. The colors and numbers denote the different cell types identified by subclustering of the major cell types. **G,** feature blots of NOX4 and **H,** violin plots for relative expression of NOX4, LUM and CNN1 in each subcluster. Electively treated abdominal aortic aneurysm (eAAA; n=4) were included in the analysis. All samples represent biological replicates. **I,** Representative alpha-smooth muscle actin and fibroblast activation protein (FAP) immunohistochemistry in human AAA tissue (n=3, biological replicates). The red stars mark the positive areas. Red/brown = AEC/DAB-positive areas; blue = hemalum positive cell nuclei. Thr, Thrombus; Int, intima; Med, media; Adv, adventitia. Scale bars represent 100 µm, 500 µm, and 1000 µm, as indicated in the figure.

Pseudotime trajectory suggest that fibroblasts and NOX4 arise as AAA progresses, supporting the observed role in fibrosis (**Supplementary Fig. 6B-E**). In contrast, the high population of smooth muscle cells in early AAA disappears along the pseudotime trajectory as does NOX4 expression in those cells (**Supplementary Fig. 7B-E**). An effect that may explain the decrease in NOX4 expression with increasing AAA diameter.

In conclusion, these data suggest that myofibroblast-to-fibroblast differentiation and smooth muscle cell (de)-differentiation in AAA are associated with increased and decreased NOX4 expression, respectively.

### NOX4 is expressed in microvascular endothelial cells in human AAA

AAA rupture has been attributed to an increased *vasa vasorum* neovascularization ^20^. In our data (n=41 AAA), a greater aortic diameter was strongly associated with increased vascularization, as indicated by the number of CD31-positive vessels per mm² (**Supplementary Fig. 8A**). This enhanced neovascularisation was associated with increased eNOS (NOS3) expression only in morphological healthy control sections (*R*²=0.48). However, this correlation was diminished in eAAA (*R*²=0.25) and was essentially absent in rAAA (*R*²=0.02), suggesting a shift in endothelial cell populations. (**Supplementary Fig. 8B**). Given that eNOS expression is highly restricted to the endothelial cells of blood vessels ^21^ and the aortic *vasa vasorum* contains both microvascular and lymph capillaries ^22^, we conclude that the CD31-positive area does not reflect the number of blood vessels in AAA. Instead, the data likely indicate an increase in microvascular vessels. Consistent with this, endothelial cells identified in AAA segregated into two major subclusters, which could be annotated as a mixed population of arterial, microvascular, and venous endothelial cells (EC.1), and a distinct group of venous/lymphatic endothelial cells (EC.2) **(Fig. 5A, Supplementary Fig. 9, Supplementary Table 7).** Notably, NOX4 expression was enriched in EC.1, whereas lymphatic endothelial cells (EC.2) exhibit very low NOX4 expression **(Fig. 5A**). GO terms and KEGG analysis suggested increased metabolic activity in EC.2 (LEC), as shown in previous studies ^23^. In addition, pathways associated with the ER and Golgi apparatus were up-regulated in LEC. In contrast, endothelial cells in EC.1 showed an inflammatory or “activated” signature **(Fig. 5B, C).**

**Figure 5:**
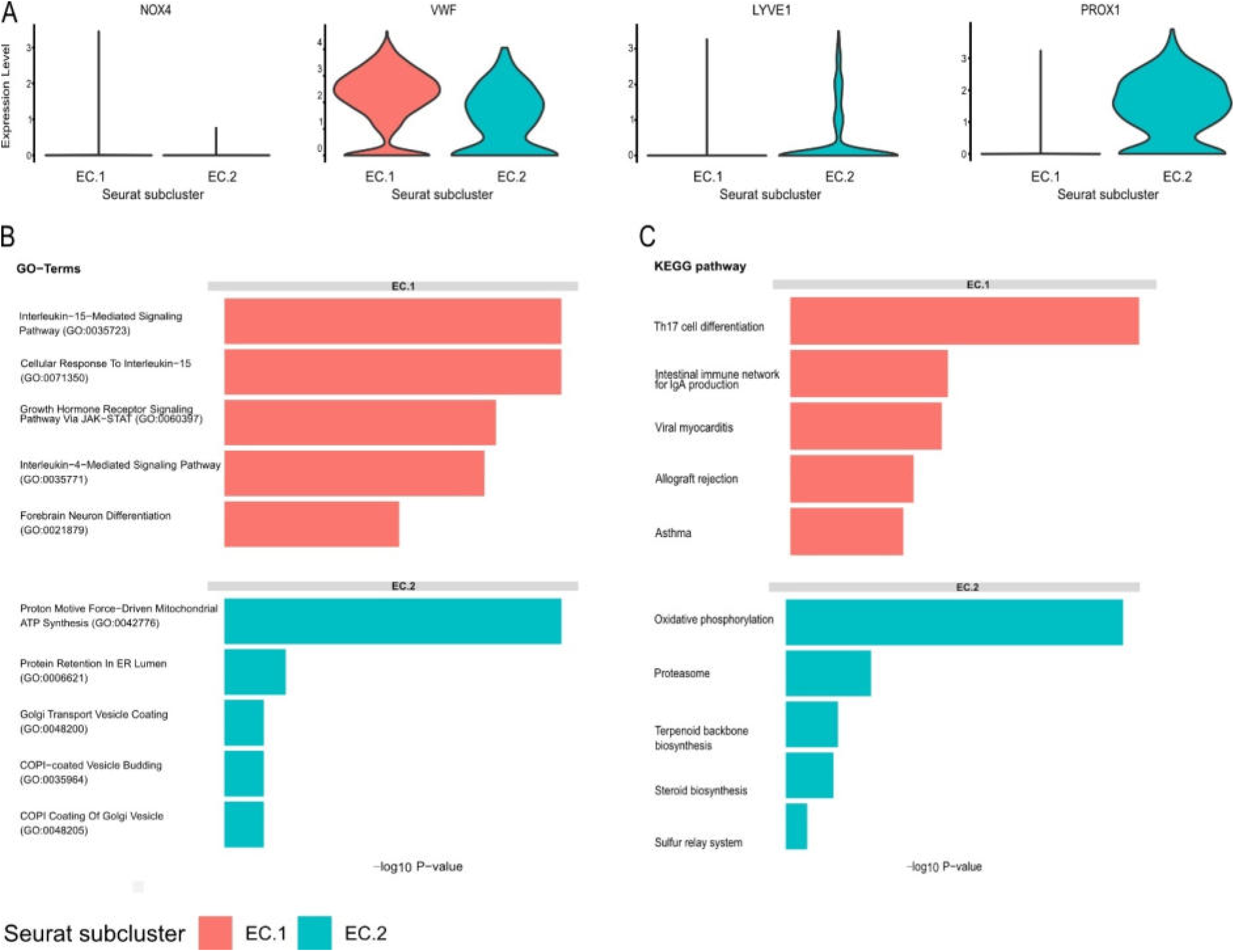
Expression of (lymph)angiogenic markers and characteristics of endothelial cells in human AAA and corresponding controls. **A,** Violin plots illustrating the distribution of manually selected genes NOX4, VWF, LYVE1, PROX1 in both endothelial cell subclusters. **B,** Top 5 up-regulated GO enrichment analysis in the two Seurat subclusters of endothelial cells. Functional analysis by GO (Top5 GO terms) for upregulated differentially expressed genes in the two indicated endothelial cell subcluster vs. the other endothelial cells subclusters. **C,** Top5 upregulated KEGG pathways in both endothelial cell subclusters. Top5 significantly enriched KEGG pathways for upregulated genes and downregulated genes in the indicated endothelial cells subcluster vs. the other endothelial cell subcluster. Electively treated abdominal aortic aneurysm (eAAA; n=4) were included in the analysis. All samples represent biological replicates.

We next investigated the transcriptional regulation underlying the observed gene expression patterns. In microvascular endothelial cells (EC.1), transcription factor analysis revealed high activity of transcription factors known to promote endothelial cell sprouting, coordinated vascular growth, ^24^ and the differentiation of mature endothelial cells from their precursors ^25^ **(Supplementray Fig. 10** and **Supplementary Table 8)**. Pseudotime trajectory analysis further suggested that microvascular endothelial cells in EC.1 may serve as precursors of lymphatic endothelial cells (EC.2), with a progressive loss of NOX4 along this transition. Notably, TGF-β1, one of the strongest inducers of NOX4 in fibroblasts and smooth muscle cells^26^ exerts its effects via TGFBR1, which displays a similar expression pattern to NOX4 (**Supplementary Fig. 11A-D**).

In conclusion, two major endothelial cell populations were identified in the vasa vasorum of late-stage AAA, highlighting a novel role for lymphatic endothelial cells. A phenotypic transition from microvascular endothelial cells to LECs was observed, accompanied by a gradual loss of NOX4 expression.

### Endothelial cell communication in human AAA associated with fibrosis and inflammation

Cell-cell communication patterns were analyzed using Cell Chat.^27^ The data strongly support a communication pathway from microvascular endothelial cells (EC.1), that express NOX4, to fibroblasts and smooth muscle cells (**Fig.6 A,B**). While lymphatic endothelial cells (EC.2) can best respond to signals from dendritic cells and monocytes/macrophages (**Fig.6 A,C**). Interestingly, both EC clusters signal via E-selectin with CD44 or golgi glycoprotein 1 (GLG1) (**Fig. 6D**). CD44 is known to modulate fibroblast function and induce fibroblast recruitment to sites of injury during tissue remodeling ^28,29^. GLG1 is a receptor for fibroblast growth factors^30^ and a negative regulator of TGF-β1 production ^31^. Lower TGF-β1 secretion could explain the reduction in NOX4, as TGF-β1 is one a potent inducer of NOX4 in vascular cells ^26^. Taken together, these data suggest that intercellular communication supports fibrotic remodeling in AAA.

**Figure 6:**
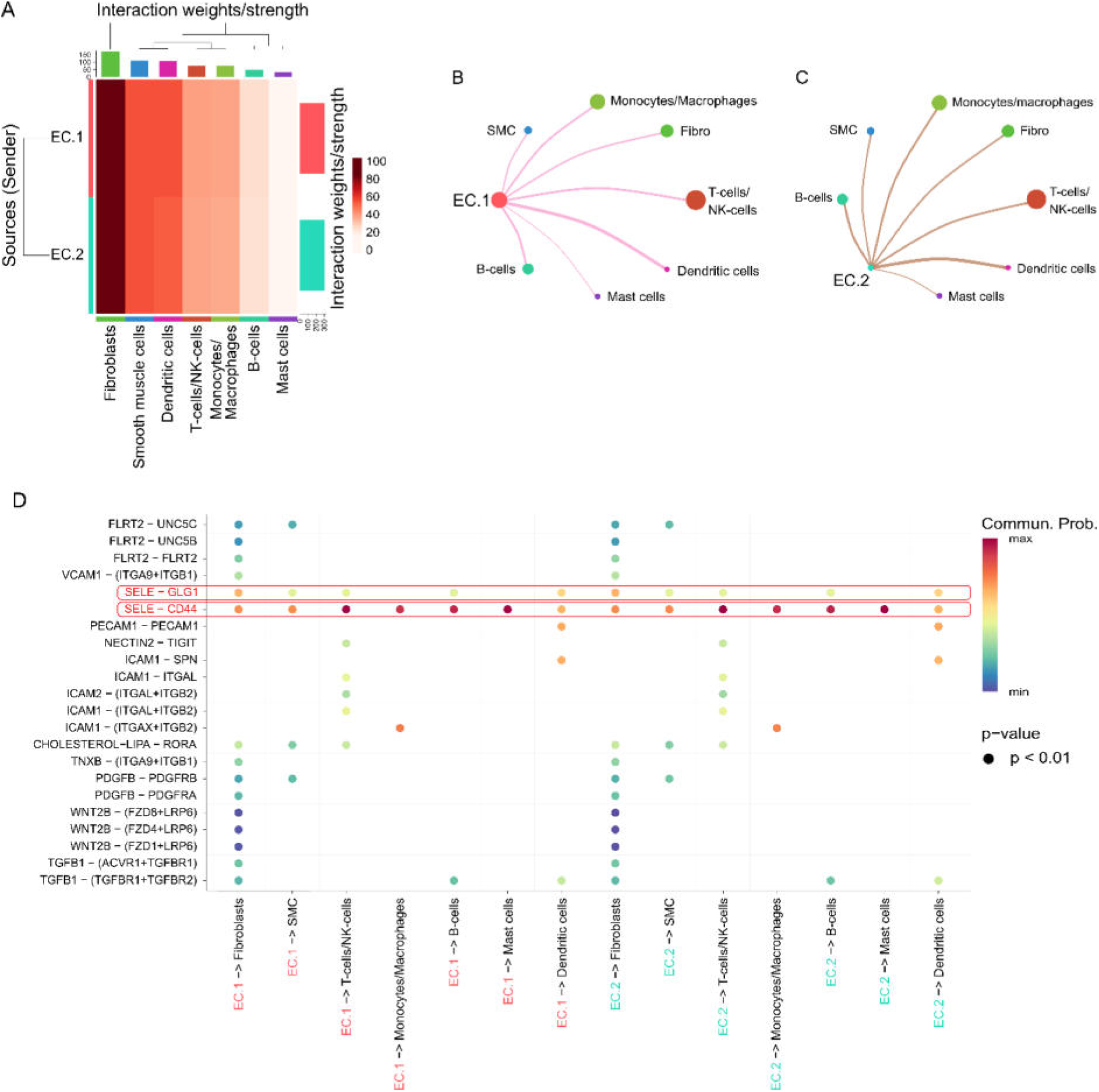
Cell-cell communication of endothelial cells in human AAA. **A,** The heat map compares the overall differential interaction of EC.1 and EC.2 as sender cells in terms of incoming signalling patterns with the receiver cells (major cell populations in AAA). The heat map shows the intensity (strength) of the interactions between cells. The darker the colour, the higher the intensity. The coloured vertical bars on the two axes represent the total number of interactions between the cell type(s). The shaded rectangles represents the strength/number of interactions between each cell type/group. Circo plots represent the interaction between different cells types. Each circle represents a different cell type/group, while the curved edges represent the interaction between the source or sender cell types (**B,** EC.1 and **C,** EC.2) and the recipient cell types. Circle sizes are proportional to the number of cells in each cell group. Edge colours are consistent with sources as senders, and edge weights are proportional to the strength of the interaction. A thicker edge line indicates a stronger signal. **D,** The bubble plot shows the relationship between EC.1 and EC.2 and the major cell populations in AAA with manually selected and ligand-receptor pairs. Colors in the bubble plot are proportional to the communication probability, where dark and yellow colors correspond to the smallest and largest values. Electively treated abdominal aortic aneurysm (eAAA; n=4) were included in the analysis. All samples represent biological replicates.

### Dynamics of small microvessls in human AAA

To further elucidate the role of (lymphatic) endothelial cells in AAA, CD31-positive ares were categorized into small (<8 µm), medium (8-26 µm) and large vessels (>26 µm) based on tertile distribution (**Fig. 7A-C**). As the aortic diameter increased, the number of small vessels progressively declined, while the proportion of large vessels increased and eventually surpassed that of small vessels (**Fig. 7D,E**). Importantly, aneurysmal NOX4 mRNA expression showed a strong positive correlation with the proportion of small vessels (**Table 2, Fig. 7D**). Moreover, the percentage of small vessels was positively associated with haemosiderin-positive areas, suggesting a link to vascular leakage (**Table 2**). Vessels less than 8 µm could be lymphatic vessels^32^, while the medium and large vessel categories likely include microvascular structures. Using single cell analysis and immunohistochemistry, we confirmed the presence of lymphangiogenesis in human AAA ^33^, as evidenced by the increased number of VEGFC, FLT4 and PROX1-expressing cells as well as expansed PROX1-positive areas in human AAA (**Supplementary Fig. 12A-D**).

**Figure 7:**
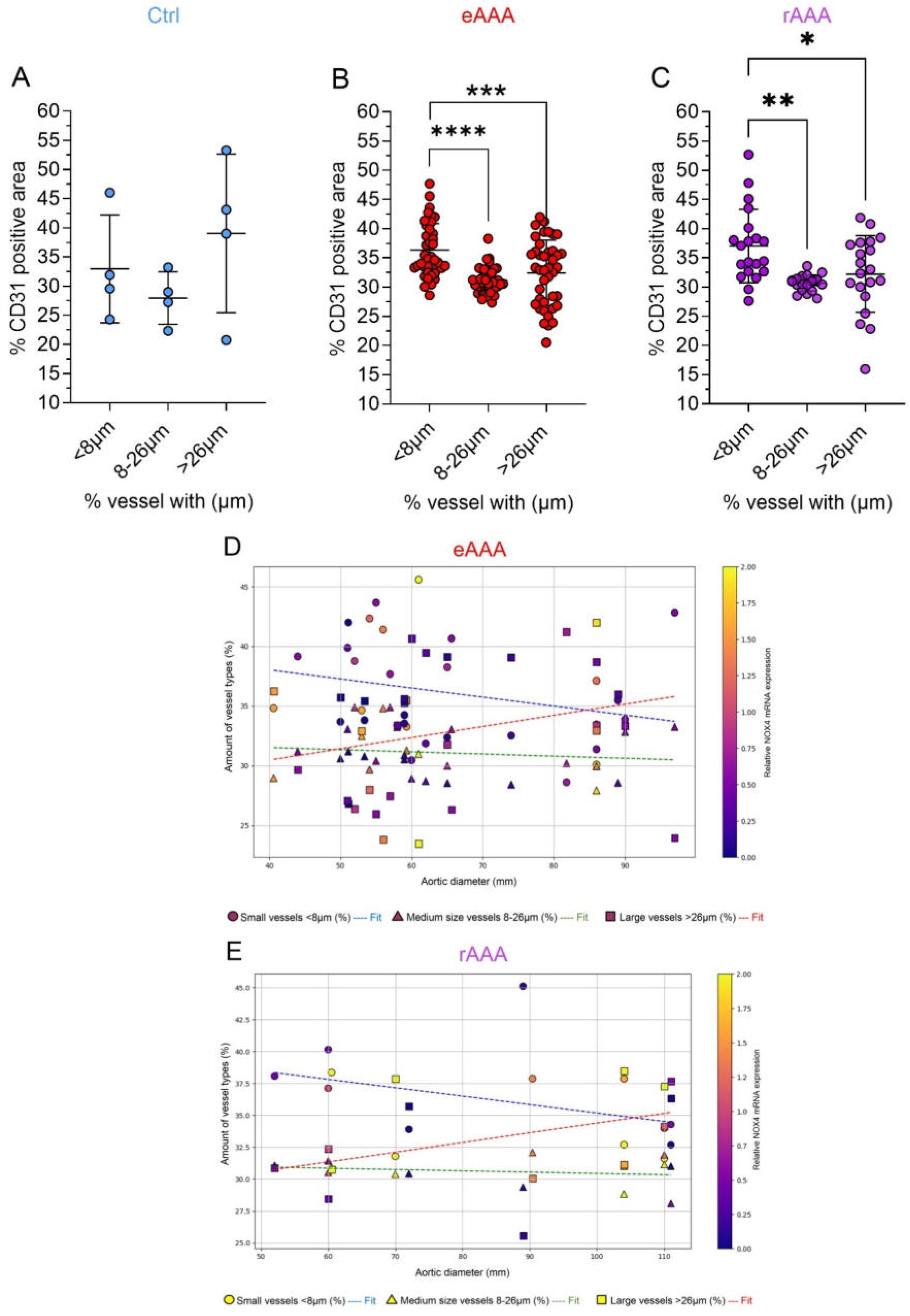
Amount of different sizes of CD31-positive vessels and associations with the aortic diameter and NOX4 mRNA expression in human AAA. Human AAA were fixed in formalin and parrafin sections were prepared. Endothelial cells were stained with CD31 using immunohistochemistry and the CD31-positive area was quantified using a macro in Image J. Tertiles were formed from all measured values and these were divided into vessels with a diameter <8 µm, 8-26 µm and >26 µm. Comparison of the percentage of CD31-positive vessels with <8 µm, 8-26 µm and >26 µm diameters in aortas from patients with **A,** non-diseased controls n=4, **B,** elevtively treated AAA (eAAA) n=49 and **C,** ruptured AAA (rAAA) n=19. Ordinary one-way ANOVA and Tukey’s multiple comparisons test. \**P*<0.05, \*\**P*<0.001.\*\*\**P*<0.0001, \*\*\*\**P*<0.00001. Associations between aortic diameter, percentage of small, medium and large CD31-positive vessels and NOX4 mRNA expression in aortas from patients with **D,** elevtively treated AAA (eAAA) and **E,** ruptured AAA (rAAA). NOX4 mRNA expression was scaled with yellow representing the highest expression and blue the lowest. **D-E,** Abdominal aortic aneurysm (eAAA; n=49), and ruptured AAA (rAAA; n=19) samples were included in the analysis. All samples represent biological replicates.

**Table 1.** Correlations between the mRNA expression of the lymphatic marker FLT4 and morphological and histopathological features of human AAA. FLT4 mRNA expression was analyzed by qPCR, the collagen content in picrosirius red stained AAA sections and MMP9 activity by zymography. Electively treated abdominal aortic aneurysm (eAAA; n≥17) were included in the analysis. All samples represent biological replicates.

|  | **FLT4 mRNA expression** |
| --- | --- |
| **Aortic diameter, mm** | r= -0.2368<br>p= 0.2162<br>n= 29 |
| **AAA volume** | r= -0.4429<br>p= 0.039<br>n= 22 |
| **Intima+Media thickness** | r= -0.500<br>p= 0.043<br>n= 17 |
| **Collagen content** | r= 0.4828<br>p= 0.05<br>n= 17 |
| **MMP9 activity** | r= -0.5273<br>p= 0.0169<br>n= 20 |

**Table 2.** Properties of small and large CD31-positive vessels in human AAA. Human AAA were fixed and paraffin sections were prepared. These were stained for the presence of Fe3+ using Perl’s Prussian blue and for the macrophage marker CD68 using immunohistochemistry. Carbonyl groups in side chains of proteins were derivatized with 2,4-dinitrophenylhydrazine (DNPH) to 2,4-dintrophenylhydrazone (DNP-hydrazone). DNP-derivatized proteins were separated on Bis-Tris protein gels and were detected with an anti-DNP antibody. Protein expression of phosphorylated and total p38MAPK, SMAD3 and JNK was detected by Western blot. The DNA binding activity of the transcription factor NF-κB was assessed by the TransAM NF-κB DNA-binding ELISA using nuclear extracts from AAA tissues. Electively treated abdominal aortic aneurysm (eAAA; n≥8) were included in the analysis. All samples represent biological replicates.

| | % of vessels <8 μm | % of vessels >26 μm |
| --- | --- | --- |
| **Fe<sup>3+</sup> positive area** | r=0.8692<br>p=0.0023<br>n=10 | r=-0.7566<br>p=0.0157<br>n=10 |
| **Carbonly residues in oxidized proteins** | r=0.8787<br>p=0.0032<br>n=9 | r=-0.6778<br>p=0.05<br>n=9 |
| **CD68 positive area** | r=0.5449<br>p=0.0018<br>n=30 | r=-0.4514<br>p=0.0123<br>n=30 |
| **p-p38/total p38 protein expression** | r=0.4522<br>p=0.092<br>n=15 | r=-0.4826<br>p=0.0702<br>n=15 |
| **p-Smad3/total Smad3 protein expression** | r=0.7665<br>p=0.0323<br>n=8 | r=-0.4762<br>p=0.0163<br>n=8 |
| **p-JNK/total JNK protein expression** | r=0.5308<br>p=0.0440<br>n=15 | r=-0.5952<br>p=0.0214<br>n=15 |
| **NF-κB binding activity expression** | r=0.5636<br>p=0.0963<br>n=10 | r=-0.6970<br>p=0.0306<br>n=10 |
| **NOX4 mRNA expression** | r=0.3912<br>p=0.03<br>n=32 | r=-0.3200<br>p=0.07<br>n=32 |

Finally, we sought to investigate a potential link between NOX4, lymphatic endothelial cells, and fibrosis in AAA. Pseudotime analysis of endothelial and fibroblast clusters in our dataset revealed a trajectory suggesting a transition from endothelial cells to fibroblasts and subsequently to myofibroblasts (**Fig. 8A,B**). Notably, a fibroblast subpopulation (Fibro.4), which retained the expression of several endothelial cell-specific genes, may represent cells in an intermediate or transitional state (**Supplementary Fig. 6A**). We observed an increase in NOX4 expression during the transition from LECs to fibroblasts, supporting our hypothesis that NOX4 plays a key role in myofibroblast formation and fibrosis^34^ (**Fig. 8C,D**). This process may be attributed to endothelial-to-mesenchymal transition (EndMT), a key mechanism in the development of fibrotic disorders, characterized by the loss of endothelial markers and the acquisition of mesenchymal features^35^.

**Figure 8:**
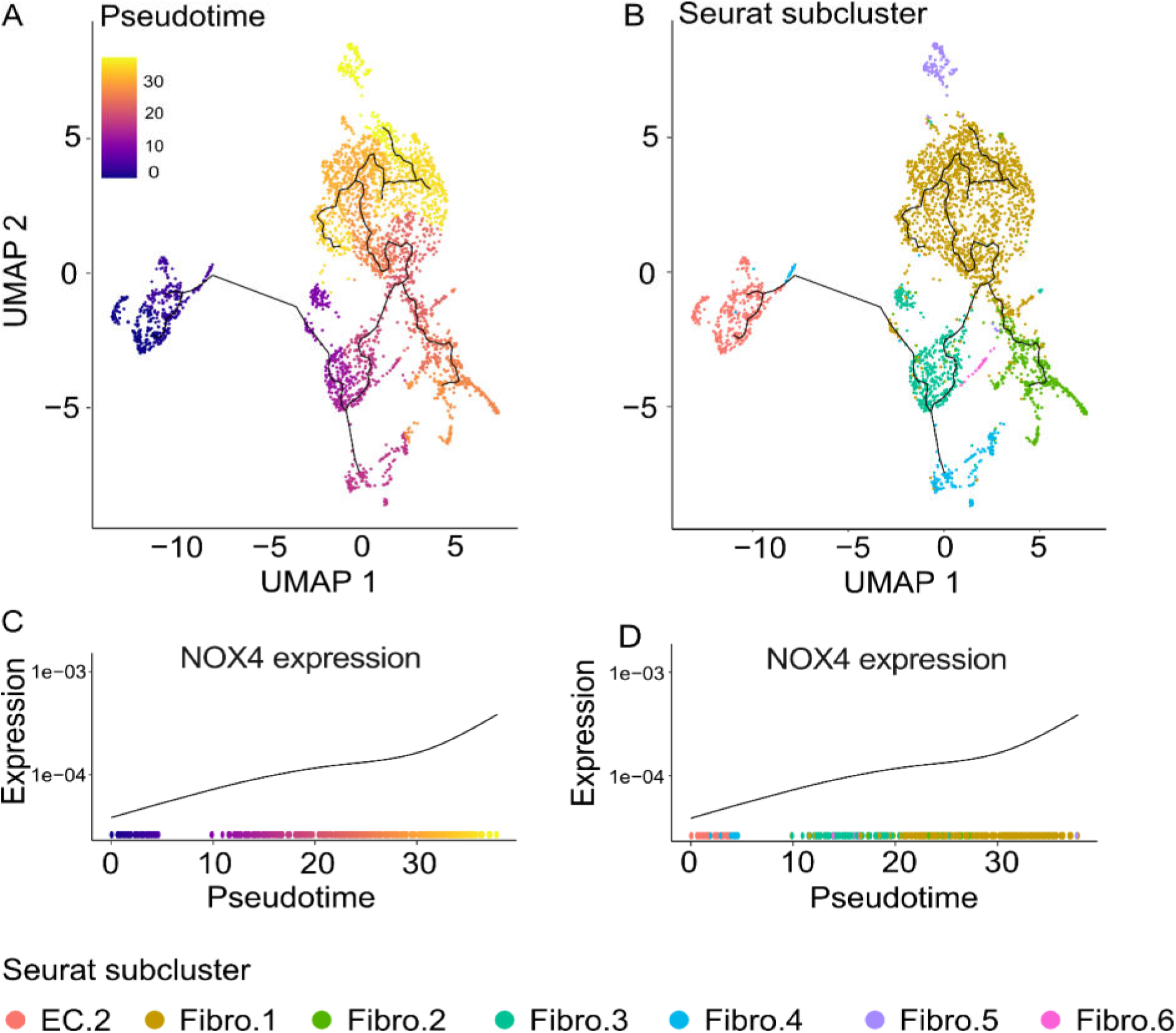
Pseudotime differentiation trajectory from fibroblasts to endothelial cells and NOX4 kinetics in human AAA. Black lines on the UMAP plots represent the trajectory graph. The root of the pseudotime trajectory is marked as “1” and was manually selected by high ACTA2 expression as previously shown for smooth muscle cells. UMAP plots with single-cell trajectories coloured by **A,** pseudotime and by **B,** Seurat supervised cell clusters. Expression of NOX4 by **C,** pseudotime and **D,** by the Seurat subclusters. Expression is based on log10. Pseudotime passess through a range of colours to represent progress through the transition. Dark blue = root cells, yellow = end cells. Cell-cell-interaction of EC.1 and EC.2 with the major cell types in AAA. Electively treated abdominal aortic aneurysm (eAAA; n=4) were included in the analysis. All samples represent biological replicates.

The data indicate changes in microvessels and lymphangiogenesis in the *vasa vasorum*. In addition, as the aortic diameter increases, the proportion of large microvessels increases.

## Discussion

In this study, we investigated the role of NOX4 in abdominal aortic aneurysm (AAA) and identified cell-specific, disease-related mechanisms that may provide new insights and opportunities needed to develop new therapeutic approaches for the treatment of AAA.

Despite the protective role of NOX4 in atherosclerosis ^12^ and general vascular inflammation ^36^, the data support a detrimental role of NOX4 in initiation and progression of AAA. NOX4 was found to force the differentiation of myofibroblasts and lyph endothelial cells. NOX4 mRNA expression was associated with pro-fibrotic effects, which consistent with a large body of literature ^37,38^. In the present study, 20 % of the patients were diabetic. It is known that type 2 diabetes leads to increased fibrosis in the vasculature due to increased ECM accumulation ^39^ and patients with carotid artery stenosis on insulin therapy have increased NOX4 expression in advanced atherosclerotic lesions ^40^.

While NOX4 contributes to endothelial homeostasis and quiescence ^41^, endothelial cells in AAA lose NOX4 and differentiate into lymphatic endothelial cells or even myofibroblasts. Aberrant lymphangiogenesis can produce poorly functioning vessels ^42^. Importantly, inflamed and activated endothelial cells promote adhesion and are associated with leukocyte diapedesis ^43^, a mechanism supported by the cell-cell interactions shown here for venous and lymphatic endothelial cells. It is possible that immune cells invade and secrete elastases, which eventually degrade the *laminae elasticae*. The detailed characterisation of the vessel walls microvessels suggest that loss of blood microvessels and lymphangiogenesis are part of the aortic wall remodeling in AAA. It might be of speculation that changes in NOX4 mRNA expression in small vessels endothelial cells may accompany or even drive leakage, inflammation and subsequent wall degradation.

The increase in endothelial cells in human AAA found in this study confirms what other studies have shown ^44^. We add that endothelial cells in AAA lose NOX4 and become lymphatic, while fibroblasts and smooth muscle cells gain NOX4 expression and contribute to fibrosis. Accelerated ‘wound healing’, if not resolved in time, leads to scarring and hence loss of contractile or elastic capacity ^45^. In the pathophysiology of AAA, stiffer and less flexible ECM may then trigger matrix degradation and inflammation, and thus worsen the disease ^46^.

In conclusion, the present study provides cell-specific insights into the role of NOX4 and different cells in end-stage human AAA. NOX4 contributes to the progression of AAA by acting mainly on differentiation at a cell-specific level. It appears that NOX4 acts differently in endothelial cells and fibroblasts, but it seems to link two important mechanisms: lymphangiogenesis and fibrosis.

AAA is a common vascular disease and rupture of an AAA is a life-threatening event. As surgical intervention is currently the only way to prevent AAA rupture, prolonging AAA progression is a valuable therapeutic target. We believe that targeting NOX4 in AAA has the potential for pharmacological intervention in AAA.

### Limitations of the present study

A limitation of the present study is the small number of AAA analysed on single cell level and the variation between individual patients. In addition, the number of cells analysed varied between AAA specimens. Despite the use of standardised protocols, we cannot completely exclude the possibility that we have obtained changes in the expression of surface markers due to slight differences in the enzymatic digestion and mechanical dissociation process. Due to the nature of late stage AAA, only a limited number of smooth muscle cells were present in each sample. Sub-clusters were defined by re-clustering cells of the same cell type using specific hallmark gene sets and defining sub-clusters by nested cuts of the hierarchical tree. This approach may retain unidentified nested subclusters. Apperantly healthy vessel sections from patients with arterial occlusive disease were used as controls in the gene expression and histopathology studies. As these patients suffer from atherosclerosis, it is possible that the controls were not 100% healthy. Furthermore, the three controls in the single cell analysis were taken from the thoracic part of the aorta. We are aware of the difference between the abdominal and thoracic parts of the aorta. However, these data were not used to draw any major conclusions and we do not have age-and sex-matched healthy controls for the abdominal region. Finally, the experimental AngII+BAPN of AAA in mice has limitations, such as the preferential manifestation of some pathologies in the thoracic part of the aorta, including aortic dissection.

## Data Availability

All data produced in the present study are available upon reasonable request to the authors

## Nonstandard Abbreviations and Acronyms

AAA: abdominal aortic aneurysm;
BAPN: ß-aminopropionitrile;
eAAA: electively treated AAA;
EC: endothelial cells;
ECM: extracellular matrix;
EndoMT: endothelial-myofibroblast transformation;
GO: Gene Ontology;
H_2_O_2_: hydrogen peroxide;
KEGG: Kyoto Encyclopedia of Genes and Genomes;
LEC: lymphatic endothelial cells;
NOX4: human NADPH oxidase 4 gene;
Nox4: murine NADPH oxidase 4 gene;
rAAA: ruptured abdominal aortic aneurysm;
SMC: smooth muscle cell;
TGF-β1: transforming growth factor-beta 1.

## Acknowledgment

The authors would like to thank Pamela Sabarstinski and Maria Walter for the excellent technical experience. 10x library preparation and sequencing was done by the Dresden-concept Genome Center at TUD Dresden University of Technology. We would like to thank Ellen Geibelt from the Light Microscopy Facility, a Core Facility of the CMCB Technology Platform at TUD Dresden University of Technology, for her support at the slide scanner. Author contributions: A.H. designed the study, A.H., B.H., C.S, F.H., M.M., I.K. and M.S.L. performed the experiments, A.H., A.S., M.M., F.H., C.S. D.M.P., M.S.L, R.A.B., A.B. and K.S. analyzed the data, M.K., A.B. and C.R. collected the samples, A.H. wrote the draft manuscript, and D.M.P., H.M., K.S. and C.R. edited the draft manuscript. All authors reviewed and approved the final manuscript.

## Literature

1. Sakalihasan N, Michel JB, Katsargyris A, Kuivaniemi H, Defraigne JO, Nchimi A, et al. Abdominal aortic aneurysms. Nat Rev Dis Primers 2018;4:34. doi: 10.1038/s41572-018-0030-7

2. Hartl F, Reeps C, Wilhelm M, Ockert S, Zimmermann A, Eckstein HH. [Open and endovascular repair of abdominal aortic aneurysms - clinical picture, evidence, results]. Dtsch Med Wochenschr 2012;137:1303–1308. doi: 10.1055/s-0032-1305055

3. Lindeman JH, Ashcroft BA, Beenakker JW, van Es M, Koekkoek NB, Prins FA, et al. Distinct defects in collagen microarchitecture underlie vessel-wall failure in advanced abdominal aneurysms and aneurysms in Marfan syndrome. Proc Natl Acad Sci U S A 2010;107:862–865. doi: 10.1073/pnas.0910312107

4. Mackay CDA, Jadli AS, Fedak PWM, Patel VB. Adventitial Fibroblasts in Aortic Aneurysm: Unraveling Pathogenic Contributions to Vascular Disease. Diagnostics (Basel) 2022;12. doi: 10.3390/diagnostics12040871

5. Davis J, Molkentin JD. Myofibroblasts: trust your heart and let fate decide. J Mol Cell Cardiol 2014;70:9–18. doi: 10.1016/j.yjmcc.2013.10.019

6. Takac I, Schröder K, Zhang L, Lardy B, Anilkumar N, Lambeth JD, et al. The E-loop is involved in hydrogen peroxide formation by the NADPH oxidase Nox4. J Biol Chem 2011;286:13304–13313. doi: 10.1074/jbc.M110.192138

7. Clempus RE, Sorescu D, Dikalova AE, Pounkova L, Jo P, Sorescu GP, et al. Nox4 is required for maintenance of the differentiated vascular smooth muscle cell phenotype. Arterioscler Thromb Vasc Biol 2007;27:42–48. doi: 10.1161/01.ATV.0000251500.94478.18

8. Yu B, Liu Z, Fu Y, Wang Y, Zhang L, Cai Z, et al. CYLD Deubiquitinates Nicotinamide Adenine Dinucleotide Phosphate Oxidase 4 Contributing to Adventitial Remodeling. Arterioscler Thromb Vasc Biol 2017;37:1698–1709. doi: 10.1161/ATVBAHA.117.309859

9. Liu Z, Luo H, Zhang L, Huang Y, Liu B, Ma K, et al. Hyperhomocysteinemia exaggerates adventitial inflammation and angiotensin II-induced abdominal aortic aneurysm in mice. Circ Res 2012;111:1261–1273. doi: 10.1161/CIRCRESAHA.112.270520

10. Yu W, Xiao L, Que Y, Li S, Chen L, Hu P, et al. Smooth muscle NADPH oxidase 4 promotes angiotensin II-induced aortic aneurysm and atherosclerosis by regulating osteopontin. Biochim Biophys Acta Mol Basis Dis 2020;1866:165912. doi: 10.1016/j.bbadis.2020.165912

11. Siu KL, Li Q, Zhang Y, Guo J, Youn JY, Du J, et al. NOX isoforms in the development of abdominal aortic aneurysm. Redox Biol 2017;11:118–125. doi: 10.1016/j.redox.2016.11.002

12. Schürmann C, Rezende F, Kruse C, Yasar Y, Löwe O, Fork C, et al. The NADPH oxidase Nox4 has anti-atherosclerotic functions. Eur. Heart J. 2015;36:3447–3456. doi: 10.1093/eurheartj/ehv460

13. Langbein H, Brunssen C, Hofmann A, Cimalla P, Brux M, Bornstein SR, et al. NADPH oxidase 4 protects against development of endothelial dysfunction and atherosclerosis in LDL receptor deficient mice. Eur Heart J 2016;37:1753–1761. doi: 10.1093/eurheartj/ehv564

14. Guzik B, Sagan A, Ludew D, Mrowiecki W, Chwala M, Bujak-Gizycka B, et al. Mechanisms of oxidative stress in human aortic aneurysms--association with clinical risk factors for atherosclerosis and disease severity. Int J Cardiol 2013;168:2389–2396. doi: 10.1016/j.ijcard.2013.01.278

15. Singh D, Rai V, Agrawal DK. Regulation of Collagen I and Collagen III in Tissue Injury and Regeneration. Cardiol Cardiovasc Med 2023;7:5–16. doi: 10.26502/fccm.92920302

16. Bruijn LE, van Stroe Gomez CG, Curci JA, Golledge J, Hamming JF, Jones GT, et al. A histopathological classification scheme for abdominal aortic aneurysm disease. JVS Vasc Sci 2021;2:260–273. doi: 10.1016/j.jvssci.2021.09.001

17. Li Y, Ren P, Dawson A, Vasquez HG, Ageedi W, Zhang C, et al. Single-Cell Transcriptome Analysis Reveals Dynamic Cell Populations and Differential Gene Expression Patterns in Control and Aneurysmal Human Aortic Tissue. Circulation 2020;142:1374–1388. doi: 10.1161/CIRCULATIONAHA.120.046528

18. Hu C, Tan H, Zhang Y, Cao G, Wu C, Lin P, et al. Fibroblast Activation Protein Acts as a Biomarker for Monitoring ECM Remodeling During Aortic Aneurysm via (68)Ga-FAPI-04 PET Imaging. Adv Sci (Weinh) 2025:e2411152. doi: 10.1002/advs.202411152

19. Lu H, Du W, Ren L, Hamblin MH, Becker RC, Chen YE, et al. Vascular Smooth Muscle Cells in Aortic Aneurysm: From Genetics to Mechanisms. J Am Heart Assoc 2021;10:e023601. doi: 10.1161/JAHA.121.023601

20. Choke E, Thompson MM, Dawson J, Wilson WR, Sayed S, Loftus IM, et al. Abdominal aortic aneurysm rupture is associated with increased medial neovascularization and overexpression of proangiogenic cytokines. Arterioscler Thromb Vasc Biol 2006;26:2077–2082. doi: 10.1161/01.ATV.0000234944.22509.f9

21. Fish JE, Matouk CC, Rachlis A, Lin S, Tai SC, D’Abreo C, et al. The expression of endothelial nitric-oxide synthase is controlled by a cell-specific histone code. J Biol Chem 2005;280:24824–24838. doi: 10.1074/jbc.M502115200

22. Sano M, Unno N, Sasaki T, Baba S, Sugisawa R, Tanaka H, et al. Topologic distributions of vasa vasorum and lymphatic vasa vasorum in the aortic adventitia--Implications for the prevalence of aortic diseases. Atherosclerosis 2016;247:127–134. doi: 10.1016/j.atherosclerosis.2016.02.007

23. Montenegro-Navarro N, Garcia-Baez C, Garcia-Caballero M. Molecular and metabolic orchestration of the lymphatic vasculature in physiology and pathology. Nat Commun 2023;14:8389. doi: 10.1038/s41467-023-44133-x

24. Winnik S, Klinkert M, Kurz H, Zoeller C, Heinke J, Wu Y, et al. HoxB5 induces endothelial sprouting in vitro and modifies intussusceptive angiogenesis in vivo involving angiopoietin-2. Cardiovasc Res 2009;83:558–565. doi: 10.1093/cvr/cvp133

25. Wu Y, Moser M, Bautch VL, Patterson C. HoxB5 is an upstream transcriptional switch for differentiation of the vascular endothelium from precursor cells. Mol Cell Biol 2003;23:5680–5691. doi: 10.1128/MCB.23.16.5680-5691.2003

26. Cucoranu I, Clempus R, Dikalova A, Phelan PJ, Ariyan S, Dikalov S, et al. NAD(P)H oxidase 4 mediates transforming growth factor-beta1-induced differentiation of cardiac fibroblasts into myofibroblasts. Circ Res 2005;97:900–907. doi: 10.1161/01.RES.0000187457.24338.3D

27. Jin S, Guerrero-Juarez CF, Zhang L, Chang I, Ramos R, Kuan CH, et al. Inference and analysis of cell-cell communication using CellChat. Nat Commun 2021;12:1088. doi: 10.1038/s41467-021-21246-9

28. Acharya PS, Majumdar S, Jacob M, Hayden J, Mrass P, Weninger W, et al. Fibroblast migration is mediated by CD44-dependent TGF beta activation. J Cell Sci 2008;121:1393–1402. doi: 10.1242/jcs.021683

29. Li S, Li C, Zhang Y, He X, Chen X, Zeng X, et al. Targeting Mechanics-Induced Fibroblast Activation through CD44-RhoA-YAP Pathway Ameliorates Crystalline Silica-Induced Silicosis. Theranostics 2019;9:4993–5008. doi: 10.7150/thno.35665

30. Zarbock A, Ley K, McEver RP, Hidalgo A. Leukocyte ligands for endothelial selectins: specialized glycoconjugates that mediate rolling and signaling under flow. Blood 2011;118:6743–6751. doi: 10.1182/blood-2011-07-343566

31. Yang T, Mendoza-Londono R, Lu H, Tao J, Li K, Keller B, et al. E-selectin ligand-1 regulates growth plate homeostasis in mice by inhibiting the intracellular processing and secretion of mature TGF-beta. J Clin Invest 2010;120:2474–2485. doi: 10.1172/JCI42150

32. Billaud M, Donnenberg VS, Ellis BW, Meyer EM, Donnenberg AD, Hill JC, et al. Classification and Functional Characterization of Vasa Vasorum-Associated Perivascular Progenitor Cells in Human Aorta. Stem Cell Reports 2017;9:292–303. doi: 10.1016/j.stemcr.2017.04.028

33. Scott DJ, Allen CJ, Honstvet CA, Hanby AM, Hammond C, Johnson AB, et al. Lymphangiogenesis in abdominal aortic aneurysm. Br J Surg 2013;100:895–903. doi: 10.1002/bjs.9128

34. Hecker L, Vittal R, Jones T, Jagirdar R, Luckhardt TR, Horowitz JC, et al. NADPH oxidase-4 mediates myofibroblast activation and fibrogenic responses to lung injury. Nat Med 2009;15:1077–1081. doi: 10.1038/nm.2005

35. Zeisberg EM, Tarnavski O, Zeisberg M, Dorfman AL, McMullen JR, Gustafsson E, et al. Endothelial-to-mesenchymal transition contributes to cardiac fibrosis. Nat Med 2007;13:952–961. doi: 10.1038/nm1613

36. Gray SP, Di Marco E, Kennedy K, Chew P, Okabe J, El-Osta A, et al. Reactive Oxygen Species Can Provide Atheroprotection via NOX4-Dependent Inhibition of Inflammation and Vascular Remodeling. Arterioscler Thromb Vasc Biol 2016;36:295–307. doi: 10.1161/ATVBAHA.115.307012

37. Hecker L, Vittal R, Jones T, Jagirdar R, Luckhardt TR, Horowitz JC, et al. NADPH oxidase-4 mediates myofibroblast activation and fibrogenic responses to lung injury. Nature medicine 2009;15:1077–1081. doi: 10.1038/nm.2005

38. Hecker L, Logsdon NJ, Kurundkar D, Kurundkar A, Bernard K, Hock T, et al. Reversal of persistent fibrosis in aging by targeting Nox4-Nrf2 redox imbalance. Sci Transl Med 2014;6:231ra247. doi: 10.1126/scitranslmed.3008182

39. Li J, Huynh P, Dai A, Wu T, Tu Y, Chow B, et al. Diabetes Reduces Severity of Aortic Aneurysms Depending on the Presence of Cell Division Autoantigen 1 (CDA1). Diabetes 2018;67:755–768. doi: 10.2337/db17-0134

40. Hofmann A, Frank F, Wolk S, Busch A, Klimova A, Sabarstinski P, et al. NOX4 mRNA correlates with plaque stability in patients with carotid artery stenosis. Redox Biol 2022;57:102473. doi: 10.1016/j.redox.2022.102473

41. Hahner F, Moll F, Warwick T, Hebchen DM, Buchmann GK, Epah J, et al. Nox4 promotes endothelial differentiation through chromatin remodeling. Redox Biol 2022;55:102381. doi: 10.1016/j.redox.2022.102381

42. Brakenhielm E, Sultan I, Alitalo K. Cardiac Lymphangiogenesis in CVDs. Arterioscler Thromb Vasc Biol 2024;44:1016–1020. doi: 10.1161/ATVBAHA.123.319572

43. Schupp JC, Adams TS, Cosme C, Jr., Raredon MSB, Yuan Y, Omote N, et al. Integrated Single-Cell Atlas of Endothelial Cells of the Human Lung. Circulation 2021;144:286–302. doi: 10.1161/CIRCULATIONAHA.120.052318

44. Holmes DR, Liao S, Parks WC, Thompson RW. Medial neovascularization in abdominal aortic aneurysms: a histopathologic marker of aneurysmal degeneration with pathophysiologic implications. J Vasc Surg 1995;21:761–771; discussion 771-762. doi: 10.1016/s0741-5214(05)80007-2

45. Frangogiannis NG. Cardiac fibrosis. Cardiovasc Res 2021;117:1450–1488. doi: 10.1093/cvr/cvaa324

46. Baxter BT, Davis VA, Minion DJ, Wang YP, Lynch TG, McManus BM. Abdominal aortic aneurysms are associated with altered matrix proteins of the nonaneurysmal aortic segments. J Vasc Surg 1994;19:797–802; discussion 803. doi: 10.1016/s0741-5214(94)70004-4

