## Supplements for "NOX4 contributes to the initiation and progression of AAA in a cell type-specific manner"

Running Title: NOX4 in human and murine AAA

Anja Hofmann[1]*#, Anupam Sinha[2]*, Christoph Schürmann[3], Bianca Hamann[1], Maria Sabater-Lleal[4,5,6], Franziska Horn[1], Marvin Kapalla[1], Margarete Müglich[1], Irakli Kopaliani[7], David M. Poitz[2], Albert Busch[1], Ralph A. Bundschuh[8,9], Henning Morawietz[10], Christian Reeps[1]*, Katrin Schröder*[3,11]

*Both authors contributed equally to the study.

[1]Division of Vascular and Endovascular Surgery, Department of Visceral, Thoracic and Vascular Surgery, University Hospital and Faculty of Medicine Carl Gustav Carus, TUD Dresden University of Technology, Germany;

[2]Institute for Clinical Chemistry and Laboratory Medicine; University Hospital and Faculty of Medicine Carl Gustav Carus, TUD Dresden University of Technology, Dresden, Germany;

[3]Institute of Cardiovascular Physiology, Medical Faculty, Goethe University, Frankfurt am Main, Germany;

[4]Unit of genomics of Complex Diseases, Institut de Recerca Sant Pau (IR SANT PAU), Sant Quintí 77-79, 17 08041 Barcelona, Spain;

[5]Cardiology Unit, Department of Medicine, Karolinska Institutet, Center for Molecular Medicine, Stockholm, Sweden

[6]Centre for Biomedical Network Research on Rare Diseases (CIBERER), Instituto de Salud Carlos III, Madrid, Spain.

[7]Department of Physiology, Medical Faculty Carl Gustav Carus, Technische Universität Dresden, Dresden, Germany;

[8]Department of Nuclear Medicine, University Hospital Carl Gustav Carus, Technical University Dresden, Dresden, Germany;

[9]Institute of Radiopharmaceutical Cancer Research, Helmholtz Zentrum Dresden-Rossendorf (HZDR), Rossendorf, Germany

[10]Division of Vascular Endothelium and Microcirculation, Department of Medicine III, University Hospital and Faculty of Medicine Carl Gustav Carus, TUD Dresden University of Technology, Dresden, Germany;

[11]German Center of Cardiovascular Research (DZHK), Partner site RheinMain, Frankfurt, Germany

**#Corresponding author:**

Anja Hofmann, PhD

Division of Vascular and Endovascular Surgery

Department of Visceral-, Thoracic and Vascular Surgery

University Hospital and Faculty of Medicine Carl Gustav Carus

TUD Dresden University of Technology

Fetscherstraße 74

D-01307 Dresden, Germany

#### Supplementary Materials and Methods

##### Abdominal Aortic Wall Collection and Ethical Approval

Aortic specimens were collected from patients undergoing elective open repair (eAAA, n=33) or surgery due to AAA rupture (rAAA, n=14) (**Supplementary Table 9**). The eAAA and rAAA group included specimens that were taken from different sites of the AAA. These biopsies were treated as individual samples without calculating the median. For this reason, the number of samples analyzed differs from the original patients included. The indicated aortic samples in the control group were obtained from patients with arterial occlusive disease (Ctrl, n=4) where the femoral or bifemoral bypass was connected to the abdominal aorta. This segment was macroscopically free of atherosclerotic lesions. For single-cell RNA-sequencing, n=4 electively treated patients were analyzed (**Supplementary Table 10**). The mean aortic diameter was assessed at the largest site of the AAA by computed tomography prior to the surgical procedure. The thickness of the intraluminal thrombus (ILT) was measured at the largest distance from the inner surface of the lumen to the outer aortic wall.

Blood lipids, C-reactive protein (CRP), risk factors, comorbidities and medical therapies were prospectively evaluated. Smoking was defined as present smoking or any kind of smoking history. Informed consent was obtained from each patient and the ethics committee of the TUD Dresden University of Technology approved this study (EK 151042017). The study is in compliance with the Declaration of Helsinki.

##### RNA Isolation, cDNA Synthesis and Quantitative Real-time PCR (qPCR) in human AAA

RNA isolation and qPCR were done as described previously.(1) Primer sequences are summarized in **Supplementary Table 11.**

##### Histology and immunohistochemistry for elastin, collagen fibers, α-smooth muscle actin, cleaved caspase-3, CD68 and CD31 and semi-quantitative assessment

Histology, immunohistochemistry and quantification of elastin degradation and positively stained areas were performed as previously described.(1) The intima-media thickness was measured using the ZEN

Blue software and the ruler function. Primary antibodies with corresponding concentrations are summarized in **Supplementary Table 12**. Representative slides are presented in each figure. The background of the representative images was automatically detected and the color composition was analyzed to balance the white balance of all images for representation. The background has been filled with white color. These changes were made only to improve the visibility of slide scanner data in the manuscript context independent of the actual data analysis.

**Staining of ferric ions ($Fe^{3+}$) by Perl's Prussian blue**

Staining of ferric ions (mainly in ferritin and hemosiderin) was performed using 10% $K_3Fe(CN)_6$ and 20% HCl in a ratio of 1:1 for 60 min at 37°C. Nuclear fast red (Carl Roth, Germany) was used for counterstaining cell nuclei. Image J quantified the positive stained blue area and data are presented in % of the total section area. The mean of two different sections from one specimen was used for data analysis.

**Protein Isolation and Western Blot**

AAA samples were homogenized in 1xRIPA buffer (10mg/100 µL) supplemented with 1:100 Halt Protease and Phosphatase Inhibitor Cocktail (Thermo Fisher Scientific, Germany) using silica beads and a Precellys 24 homogenizer (VWR, Germany). Ultrasonication was used to remove residual DNA. Protein concentration was determined using BCA Protein Assay Reagent (Thermo Fisher Scientific, Germany). Proteins (15-30 µg) were separated on 4-12 % Bis-Tris protein gels (Thermo Fisher Scientific, Germany) and transferred to nitrocellulose membranes. The membranes were incubated with primary antibodies against phospho-Smad3 $Ser^{423/425}$, phospho-Akt $Ser^{473}$, phospho-p38 $MAPK^{Thr180/Tyr182}$ and total SMAD3, total p38MAPK, phospho-c-Jun $NH_2$-terminal kinase (JNK) and total-JNK(1). Primary antibodies and their corresponding molecular weights are summarized in **Supplemental Table 13.** Protein expression was detected using Immobilon Western HRP Substrate (Merck, Darmstadt, Germany) and quantified using Image J software. Protein expression was normalized to a 70 kDa band obtained after Ponceau S staining. For phosphorylation studies, total protein was expressed relative to Ponceau S and this quotient was multiplied by phosphorylated protein.

To ensure comparability of data, an internal control was run on each Western blot and the relative protein expression was normalized. For some patients only a small amount of tissue was available and therefore not every protein was analyzed in all samples. The number of samples analyzed is given in the figure legends.

**Quantification of Carbonyl Residues in Oxidized Proteins by the Protein Carbonyl Assay Kit**

Carbonyl groups in oxidized proteins were detected by the Protein Carbonyl Assay Kit (ab178020, Abcam) as described previously(1).

**Isolation of nuclear extracts from AAA tissue and assessment of DNA-binding activity of NF-κB transcription factor by TransAM NF-κB DNA-binding ELISA**

Nuclear extracts from AAA tissue were isolated using the Nuclear Extraction Kit (ab113474, Abcam). In brief, 1g/5 mL aortic tissue was minced with a scissor, mechanically ground with a mortar and pestle and sheared with a 15-gauge needle. Isolation of the nuclear and cytosolic fraction was performed according to the manufacturer's instructions, except that the extraction buffer step was repeated twice as additional washing steps. Protein concentrations were determined using Roti-Nanoquant (K880, Carl Roth) at a dilution of 1:20 for the nuclear extracts. The DNA-binding activity of NF-κB was determined by TransAM NFκB Chemi p65 (40097, Active Motif) according to the manufacturer's instructions. For NF-κB, 2 μg nuclear extract were used per well. Data are normalized to a positive control (=1) provided by the supplier.

**Quantification of matrix metalloprotease-9 (MMP9) and matrix metalloprotease-2 (MMP2) activity and expression of pro-forms by zymography**

From all samples analyzed for LOX-1 expression, zymography was performed as described elsewhere(1, 2). An internal control (=1) was run on every gel and data are presented relative to this control to ensure comparability of data.

**Assessment of elastin degradation in human AAA and mice aortae**

Degradation of elastic laminae was classified into a score of 1-4 (2) by eight persons blinded to the experiment. These persons were trained prior to the final scoring.

**Aortic Dissociation and Droplet-based scRNA-sequencing**

Aortic tissues were cut into small pieces and washed vigorously with cold 1xDPBS. Tissue was digested in an enzyme mix of 3 mg/mL collagenase type II (LS004176, Worthington Biochemical Corp.), 0.15 mg/mL collagenase type XI (H7657, Sigma Aldrich), 0.25 mg/mL soybean trypsin inhibitor (LS003571, Worthington Biochemical Corp.), 0.1875 mg/mL elastase (LS002292, Worthington Biochemical Corp.), 0.24 mg/mL hyaluronidase type I (H3506, Sigma Aldrich) and 2.38 mg/mL HEPES (15630-056, Gibco) dissolved in Hanks balanced salt solution (HBSS, 14175-053, Gibco) for 1 h at 37°C. Tissue dissociation was controlled using a microscope. Afterwards, the cell suspension was passed through a 40 µm cell strainer, centrifuged at 300xg for 10 min at 4°C, re-suspended in HBSS supplemented with 5% FBS and placed on ice for 30 min. After an additional washing of cells the remaining erythrocytes were lysed by adding 1 mL ACK Lysis buffer (Gibco) followed by a washing step. Dead cells were removed using the dead cell removal kit (Miltenyi Biotec) according to the manufacturer's instructions. Cells were resuspended in HBSS with 5 % FCS and 2 % RiboLock RNase inhibitor (Thermo Fisher Scientific) at a concentration of 1000 cells/µL.

**Droplet based Single-cell Transcriptomics**

Cells were carefully mixed with reverse transcription mix before loading the cells on the 10X Genomics Chromium system (3) in a Chromium Single Cell B Chip and processed further following the guidelines of the 10X Genomics user manual (v3.1). In short, the droplets were directly subjected to reverse transcription, the emulsion was broken and cDNA was purified using Silane beads. After the amplification of cDNA, the 10X Genomics single cell RNA-seq library preparation - involving fragmentation, dA-Tailing, adapter ligation and indexing PCR – was performed based on the manufacturer's protocol. After quantification, 10x libraries were sequenced on an Illumina NovaSeq 6000 in 100bp paired-end mode, targeting 25k fragments per cell.

#### Single-cell data analysis and computational analysis

##### Data preprocessing

*Library construction and sequencing*

The raw sequencing data was processed with the 'count' command of the Cell Ranger software (v6.0.1 and v6.1.2) provided by 10X Genomics. To build the reference, the human genome (hg38) as well as gene annotation (Ensembl 98) were downloaded from Ensembl and the annotation was filtered with the 'mkgtf' command of Cell Ranger (options: '—e attribute=gene_biotype:protein_coding--attribute=gene_biotype:lincRNA –attribute=gene_biotype:antisense'). Genome sequence and filtered annotation were then used as input to the 'mkref' command of Cell Ranger to build the appropriate Cell Ranger Reference. Single cells were generated from four individual AAA patients (AAA1 - AAA4). The table summarizes the analyzed cells.

| | **Estimated number of cells** | **Median Genes per cell** | **Fraction Reads in cells, %** | **Median UMI counts per Cell** |
|---|---|---|---|---|
| **AAA1** | 6,935 | 434 | 64.2 | 950 |
| **AAA2** | 280 | 56 | 46.6 | 716 |
| **AAA3** | 9,058 | 2,169 | 91.0 | 6,858 |
| **AAA4** | 8,928 | 2,376 | 93.0 | 7,340 |

##### Data Normalization, Dimensional Reduction, Clustering and Integration

Seurat (v4.3.2) (4) was used to perform "CreateSeuratObject" was run using the parameters: min.cells = 3, min.features = 200, to create four Seurat objects, one for each of the patient samples. Subsequently, only the cells with less than 10% mitochondrial content were retained for further analyses. This resulted in 5332, 54, 8507, 8362 cells in AAA1, AAA2, AAA3 and AAA4 respectively. Control samples (ID: GSE155468) were downloaded from GEO. Samples GSM4704932, GSM4704931 and GSM4704933 (henceforth denoted as control1, control2 and control3) were chosen for further analyses. These samples were pre-processed as described above.

For further downstream analyses of the AAA samples, it was imperative to create a unified data container in the form of a Seurat Object. For this, we first ran the “merge” command to create a consolidated Seurat object “all_AAA” of all the patients. Subsequently, data scaling, normalization and variance stabilization was done by running “SCTransform” command on “all_AAA” using the parameters: *vst.flavor= "v2",ncells=10000,variable.features.n = 10000.*

All control samples were subjected to the same procedure to create an “all_Ctrl” Seurat object. Subsequently, AAA and control samples merged together to create a third separate Seurat Object called “all_AAA_all_Ctrl”.

Dimension reduction was performed using “RunPCA” with parameter *npcs = 50.* Subsequently, “RunUMAP” and “FindNeighbors” commands were run using the parameters: *reduction = "pca", dims = 1:50.* Finally, “FindClusters” method was executed by setting *resolution = 0.1* and *algorithm=1*(original Louvain algorithm).

The data were also subjected to the alternate normalization and scaling pipeline by running commands “NormalizeData” (with parameters: *normalization.method=”LogNormalize”, scale.factor=10000*), “FindVariableFeatures” (with parameters: *selection.method= “vst”, features =10000*) and “ScaleData” (with parameters: *vars.to.regress=c(“nCount_RNA”,”percent.mt”)*) sequentially.

Visualization of the data was performed by nonlinear reduction using the method of uniform approximation and projection (UMAP) in two dimensions.

**Cell Cluster Annotation**

Cell clusters were annotated in a supervised manner using the cell-specific markers extracted from literature. Briefly, expressions of marker genes were projected onto the clusters derived from previous steps. Subsequently, visualizations of marker genes were performed using violin plots. The marker genes whose mapping led to the most unique separation of clusters were retained and the clusters annotated with cell-type definition.

**Trajectory Analysis**

scRNA-seq data, even when generated from a single time point, can be used to decipher the underlying gene expression dynamics of the cardinal biochemical processes. This is essentially because the cells in scRNA-seq data are generally derived from continuous biological processes and have gradually varying transcriptomes. Pseudotemporal trajectory construction positions cells along a trajectory on the basis of their transcriptome variability. Pseudotime analysis, as it is generally called, was performed using monocle3. The "RNA" assays of Seurat objects were extracted and used as inputs. The root for fibroblasts and smooth muscle cells (SMCs) was manually set based on its high ACTA2 expression. For the two endothelial cell (EC) Seurat subclusters (EC0, EC1) the root was classified as EC0 based on the low expression of the lymphatic EC marker PROX1. A principal graph denoting the pseudotime trajectory was constructed using the "learn_graph" command with parameters: *use_partition=FALSE, close_loop=FALSE*. The chosen parameter values ensured that only one trajectory connected all the cell clusters together instead of each cell-cluster having its own trajectory. In addition, the constructed graph did not loop, as we did not expect the AAA disease progression to be a cyclical process. To order the cells, the roots (either representing the normal/non-AAA cells or the AAA cells) of the trajectories were chosen using biologically meaningful information in the form of expression of select genes (that are known to reflect the status of cells under consideration). The rest of the cells represent the progression along the AAA disease trajectory. The progress through the pseudotime trajectory was depicted using UMAP plots (colored by pseudotime, clusters, or expression of marker genes).

**Differential Gene Expression and Functional Pathway Analysis**

FindMarkers command of Seurat was used to perform differential gene expression analyses (between two different clusters or conditions) with the parameter logfc.threshold set to 0.1. Similarly, FindAllMarkers command was used to perform differential gene expression analyses (between all the clusters) with the parameter logfc.threshold set to 0.1. Wilcoxon rank sum test and false discovery rate (FDR)/Benjamini-Hochberg correction were used for hypothesis testing. enrichR(5) was run on differentially expressed genes using the KEGG and GO databases.

**Transcription Factor Binding and Regulon Activity**

To categorize Cell-States/Types we calculated Transcription Factor Activity (TFA) using decoupleR(6), an R package. TFA is calculated using the expression of genes under the transcriptional control of the Transcription Factor (TF) in question. Since TFA is based on a set of genes regulated by a TF (regulons), it is useful in scoring the Cell-States of scRNA-seq data at a much finer resolution. The count data was used as one of the inputs for the "run_ulm" command from decoupleR. The other input was the regulon information extracted using the get_collectri command. The TFA values were subsequently normalized, scaled and used for plotting.

**Cell-cell-communication ligand-receptor interactions**

CellChat, an R package was used to perform cell-cell communication analyses. Data and annotations from Seurat objects were used for creation of CellChat object using the command "createCellChat" . For the calculations, the Endothelial Cell clusters were used as sources/senders of the signals and the immune cells were used as receivers. Subsequently, the commands netVisual_circle, netVisual_heatmap, netVisual_bubble and plotGeneExpression were used for plotting.

**Bulk RNA Sequencing**

Differential gene expression in AAA and healthy controls was performed using bulk RNA-seq data on 96 AAA aortas and 44 control aortas from deceased donors. Detailed methods about subject collection, sample processing and RNA sequencing have been described elsewhere(7). In brief, data was created using a read length of 150 bp and paired-end sequencing. We used STAR v.2.5.3(8) to perform the alignment on the reference genome version GRCh38 and we then used RSEM v1.3.0(9) for gene quantification. Gene quantifications were expressed as Transcripts Per Million (TPMs), which were obtained by normalizing for gene length first, and then for sequencing depth. Before conducting differential expression analyses, we normalized the TPMs counts using quantile normalization and removed lowly expressed genes with less than 0.5 TPMs in more than 50% of the samples. We calculated differential expressed genes between AAA and controls using linear regression. All comparisons were adjusted for age and sex and the identified technical covariates, and genes known to be affected by ischemic time were removed from the significantly differentially expressed genes list.

**Comparison with non-AAA Control Samples**

Because aortic segments from age-matched patients without aneurysmal disease were not available, the data presented here were compared with sequencing data from a previously published study on segments of the ascending aorta.(10) Sequencing data were used for trajectory interference and transcription factor enrichment analysis.

**Nox4 Knockout Mice**

Mice were housed in a 12/12 day and night cycle with unlimited access to food and water. All animal experiments were performed in accordance with the National Institutes of Health Guidelines on the Use of Laboratory Animals. The University Animal Care Committee and the Federal Authorities for Animal Research (Darmstadt, Germany) approved the study protocol (TVG FU1096). Nox4-/- mice were generated by targeted deletion of the translation initiation site and of exons 1 and 2 of the Nox4 gene2 and backcrossed into C57/Bl6J for 5 generations. Breeding was carried out by crossing heterozygous animals so that knockouts and direct wild-type littermates controls (WT) could be used throughout the studies.(11) C57Bl6J mice served as controls and were purchased from Janvier (Le Genest Saint Isle, France).

**Angiotensin II and β-Aminopropionitrile-induced (BAPN)-induced AAA Formation**

Male mice at 8-10 weeks of age were used. AAA were induced by systemic infusion of Ang II (#4006473, Bachem) in combination with an atherogenic diet (D12336, Research Diets Inc) for 4 weeks

(28 days). Osmotic pumps (model 2001, Cupertino, USA) filled with AngII were implanted subcutaneously into the mice. For the procedure, animals were once anesthetized via nasal administration of 2–3% isoflurane. Heart rate was maintained between 450 and 500 bpm, and body temperature was continuously monitored and controlled throughout the surgery. The osmotic pump released Ang II consistently to achieve a dose of 1 mg/kg/d. AAA progression was additionally forced by ß-aminopropionitrile (BAPN) administration in the drinking water at a dose of 1 mg/mL at the first 2 weeks of treatment. BAPN is a lysyl oxidase inhibitor that blocks the cross-linking of collagen and elastin and thereby weakens the aortic wall.(12) The AngII plus BAPN mouse model has been shown to induce morphological and histological characteristics similar to human AAA without using hyperlipidemic backgrounds and thereby enhancing atherosclerosis.(13) This model was chosen because it allowed us to simulate the human findings of NOX4 and fibrosis in a more appropriate way. Blood pressure was measured on five consecutive days in conscious mice using a non-invasive tail-cuff method with a 6-channel setup (Visitech Systems, NC). Development of AAA was monitored by analyzing the aortic diameter at maximal dilation using high-resolution 2-dimensional ultrasound imaging (B-mode) with a Vevo 3100 ultrasound machine (Toronto, Canada). Mice were sacrificed directly after the 28 days treatment period and the entire aorta was removed at segments with the indicated distance from the right common carotid artery (RCCA). AAA was confirmed as a dilated segment of the suprarenal aorta and by histological evaluation. Rupture of the thoracic aorta or abdominal aorta as the cause of death was defined by the presence of a hematoma/blood clots in the thoracic or abdominal cavities of these mice.

**Histology of Aortic Sections**

Aortic sections were fixed in 4 % PFA, dehydrated in 30 % sucrose and embedded in Tissue-Tek O.C.T. Sections of 10 µm were from the RCCA to the aortic bifurcation. Histological slides were stained with Sirius red and hematoxylin and eosin. The lumen (maximum external aortic diameter) was determined by planimetry in HE-stained sections using the ImageJ program. Collagen density was measured in Sirius-red stained sections using polarized light microscopy.

**Statistical Analysis**

Grubb's test was used to detect significant outliers in all data sets. Outliers within each group are indicated in each figure legend. Normality was tested by the D'Agostino and Pearson normality test. Non-Gaussian distributed data were analyzed by Mann Whitney U or Kruskal-Wallis and Dunn's multiple comparisons test and Gaussian distributed data by unpaired t-test or One-Way Analysis of Variance (ANOVA) and Holm-Šídák's multiple comparisons test, respectively. Correlational analysis in non-Gaussian-distributed data was done using Spearman's correlation coefficient ($r_S$), Gaussian distributed data was compared by Pearson's correlation coefficient ($r_P$). Data are plotted as scatter dot plots, and the horizontal line shows the median with range. Differences in the distribution of cardiovascular risk factors and medical therapies across the three independent groups (Ctrl, eAAA, rAAA) were compared by Fisher's exact test using a contingency table. The null hypothesis ($H_0$) postulated that distributions of risk factors and medical therapies are independent of the outcome of the disease (Ctrl, eAAA or rAAA) and no differences will be observed between all groups. Graph Pad Prism 10.0 (GraphPad Software, Inc., La Jolla, CA, USA) software was used for statistical analysis and $P<.05$ was considered as significant. The correlational data (Figure 7, Supplementary Figure 8) were analyzed using python 3.12.9. in a Jupiter notebook with pandas, numpy and matplotlib.pyplot. The code is available upon request.

### Supplementary Figures

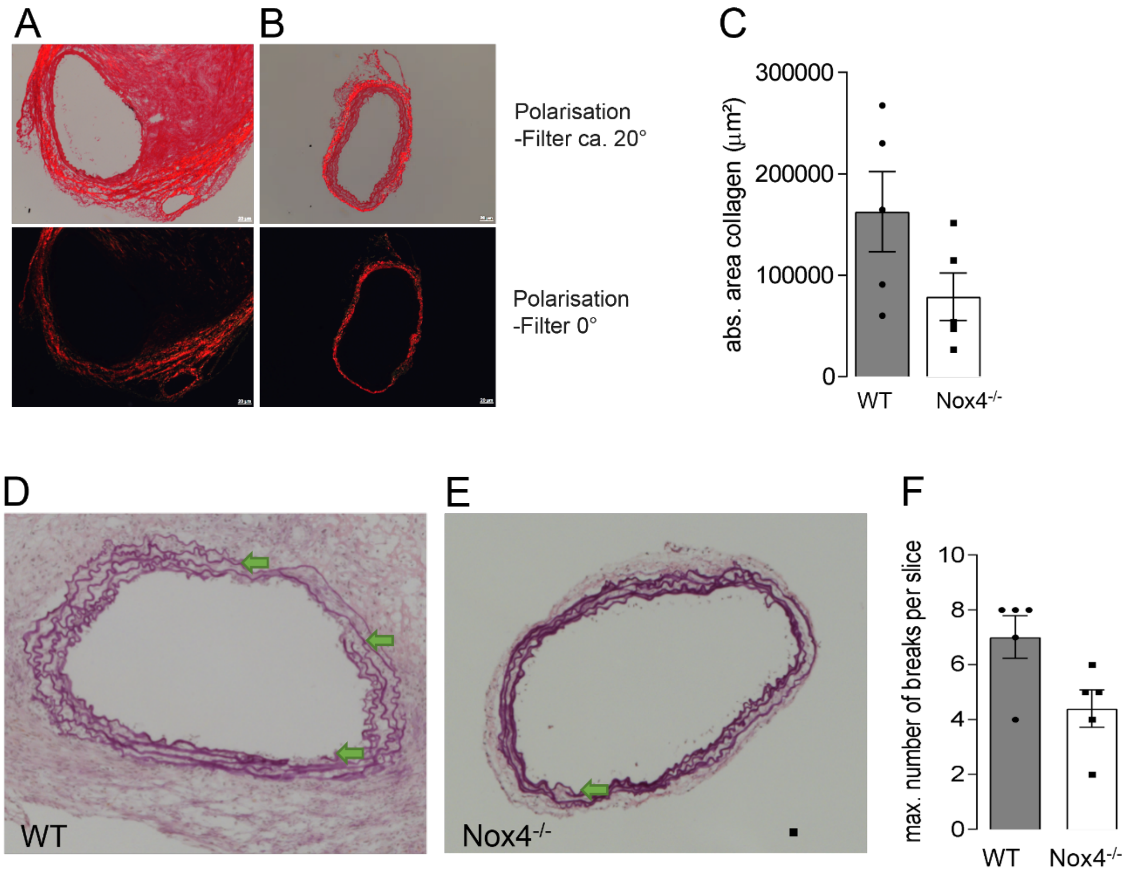


**Supplementary Figure 1: Nox4 deletion limits disruption of aortic elastic fibers and adventitial collagen.** WT and Nox4−/− mice were treated with a combination of AngII (1 mg/kg/day) and a high-fat diet for 4 weeks. AAA formation was additionally induced by BAPN administration in the drinking water (1 mg/mL) during the first 2 weeks of treatment. Representative images of aortic cross sections with stained for collagen (red) in **A,** WT and **B,** Nox4−/− mice and **C,** average of all strand breaks throughout the aorta. Representative images of aortic cross sections with arrows (green) indicating elastic fiber breaks in **D,** WT and **E,** Nox4−/− mice. **F,** Average of all strand breaks throughout the aorta. n=5 animals per group. Scale bar: 20 µm.

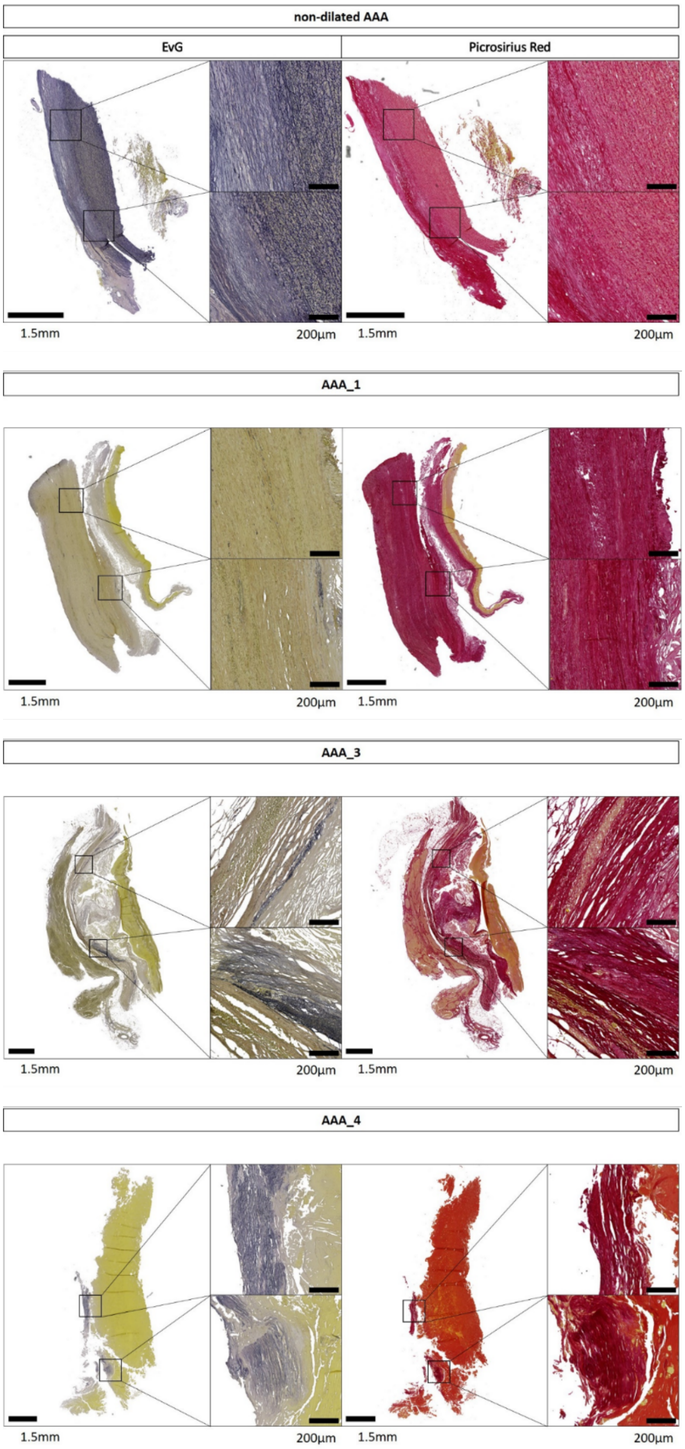


**Supplementary Figure 2: Representative slides of Elastica-van Gieson and Picrosirius red staining in n=3 aortic samples from patients with AAA used for single-cell RNA-sequencing.** A specimen from the non-dilated zone (n=1) of the abdominal aorta served as a reference. Due to material limitations, no histology of AAA_2 was available. AAA_1, AAA_3, AAA_4: n=3 biological replicates. Scale bar left picture: 1.5 mm and right picture: 200 µm.

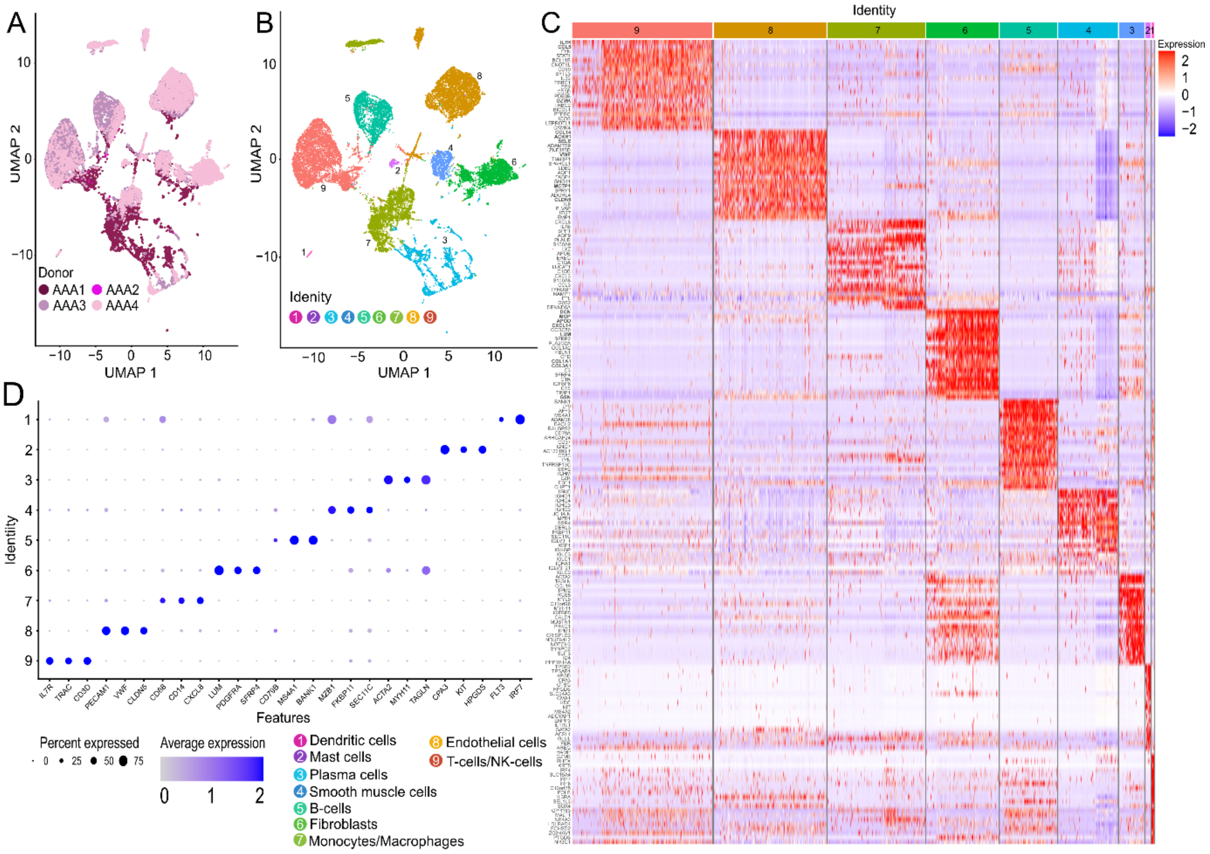


**Supplementary Figure 3: Identification of cell partitions, cell-type labeling and differential gene expression analysis in human AAA. A,** Uniform Manifold Approximation and Projection (UMAP) plot of cells from four different patients with AAA (AAA_1-AAA_4). The colors indicate the different AAA samples. **B,** UMAP plot of the different cell clusters within each AAA. The colours and numbers indicate the different cell types identified by the expression of classical marker genes. Notably, expression of natural killer cells markers (CD7, KLRC1, GNLY, KLRD1, PRF1) was detected within the T-cell cluster, but no single NK-cell population was identified and the cluster was termed NK-cells/T-cell/T-cells. Furthermore, plasma cell markers (SEC11C, PRDX4, SSR4, MZB1) were also found within all other cell clusters, but their expression remained highest in the plasma cell cluster. **C,** Dotplot showing cell type marker genes leading to the identification of monocytes/macrophages, ECs, VSMCs, fibroblast, NK-cells/T-cells, T- and B-Lymphocytes, mast cells, dendritic cells and plasma cells. The size of the dots indicates the proportion of cells expressing each gene and the dot colour of the dots indicates the expression of each gene. **D,** Cluster-defining heat map of the top 20 most enriched genes with specific expression for each cell cluster in all integrated AAA ordered by log2 fold changes (scale). Colour scheme is based on relative gene expression (z-score) and comparison of the selected cluster with all others. Electively treated abdominal aortic aneurysm (eAAA; n=4) were included in the analysis. All samples represent biological replicates.

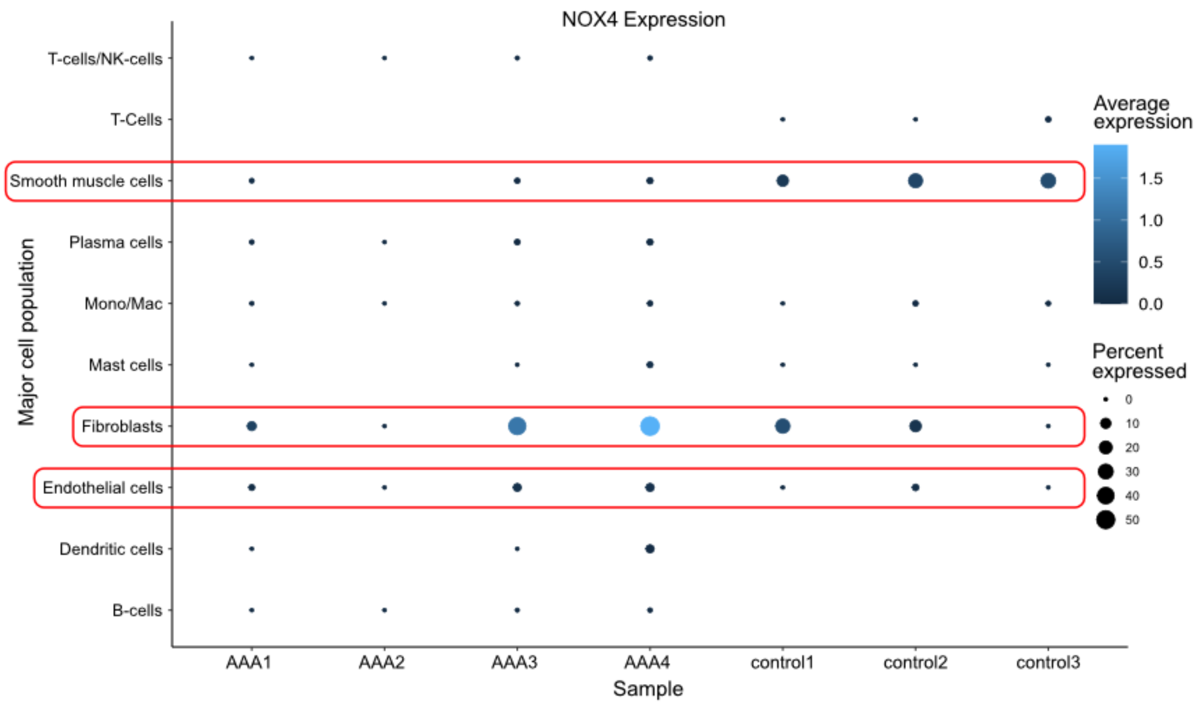


**Supplementary Figure 4: Expression of NOX4 in major cell types in human AAA and corresponding non-AAA controls.** Feature plot of relative expression of NOX4 in **A,** AAA1-2 and **B,** non-AAA controls (control1-control3). Electively treated abdominal aortic aneurysm (eAAA; n=4) were included in the analysis. All samples represent biological replicates.

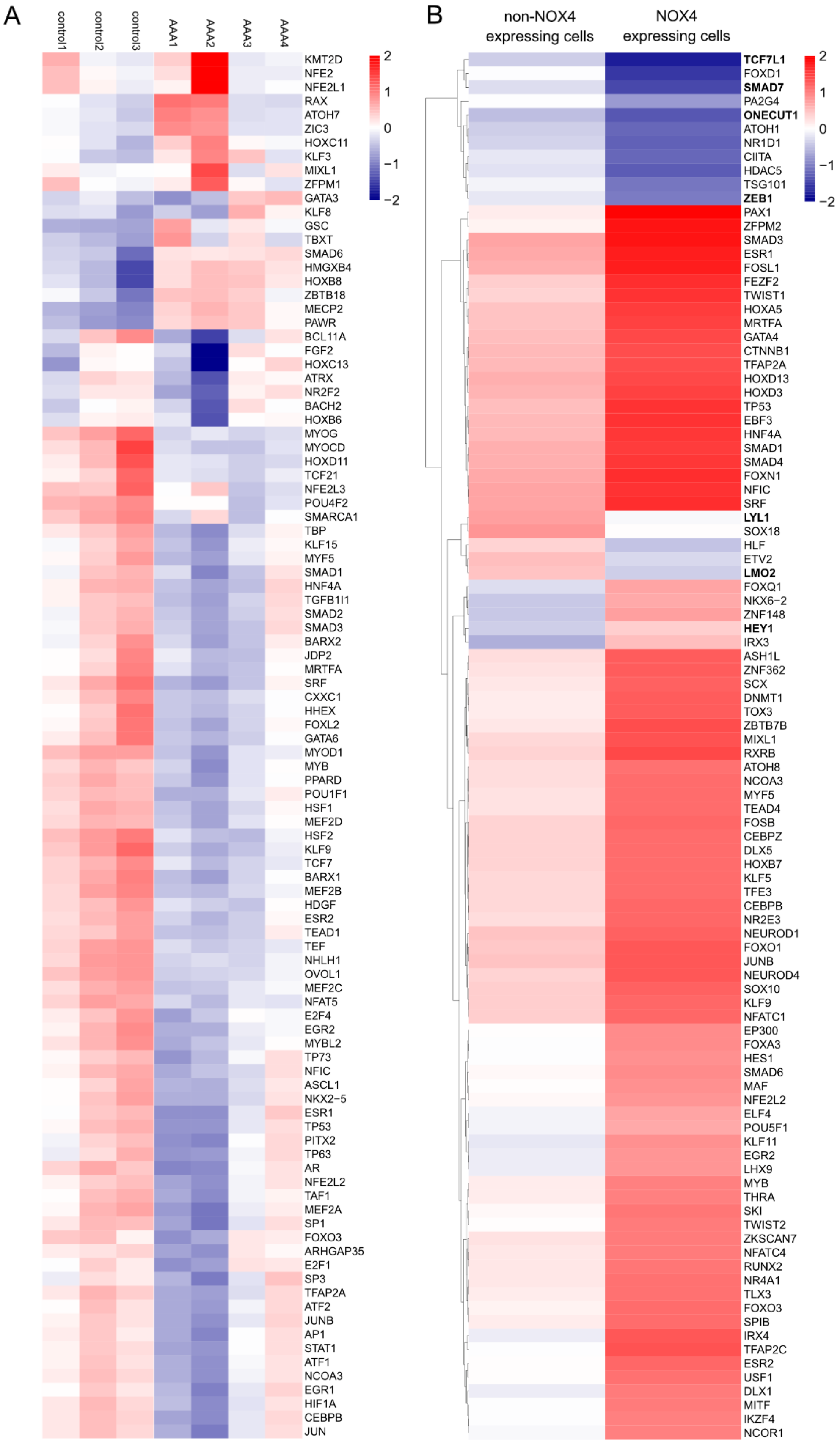


**Supplementary Figure 5: Prediction of transcription factor activities in AAA and non-diseased controls and in NOX4-expressing cells in human AAA. A,** Heatmap visualizing the significantly differentially active transcription factors compared between AAA1-AAA4 and non-diseased controls (open source data). **B,** Heatmap visualizing the significantly differentially active transcription factors between NOX4-expressing and non-NOX4-expressing cells. The heatmaps were generated by aggregated scATAC-seq data as input and the predicted regulon activity and transcription factor gene expression are shown. Colors show TF with high (z-score = 2) and low activity (z-score = -2). Electively treated abdominal aortic aneurysm (eAAA; n=4) were included in the analysis. All samples represent biological replicates.

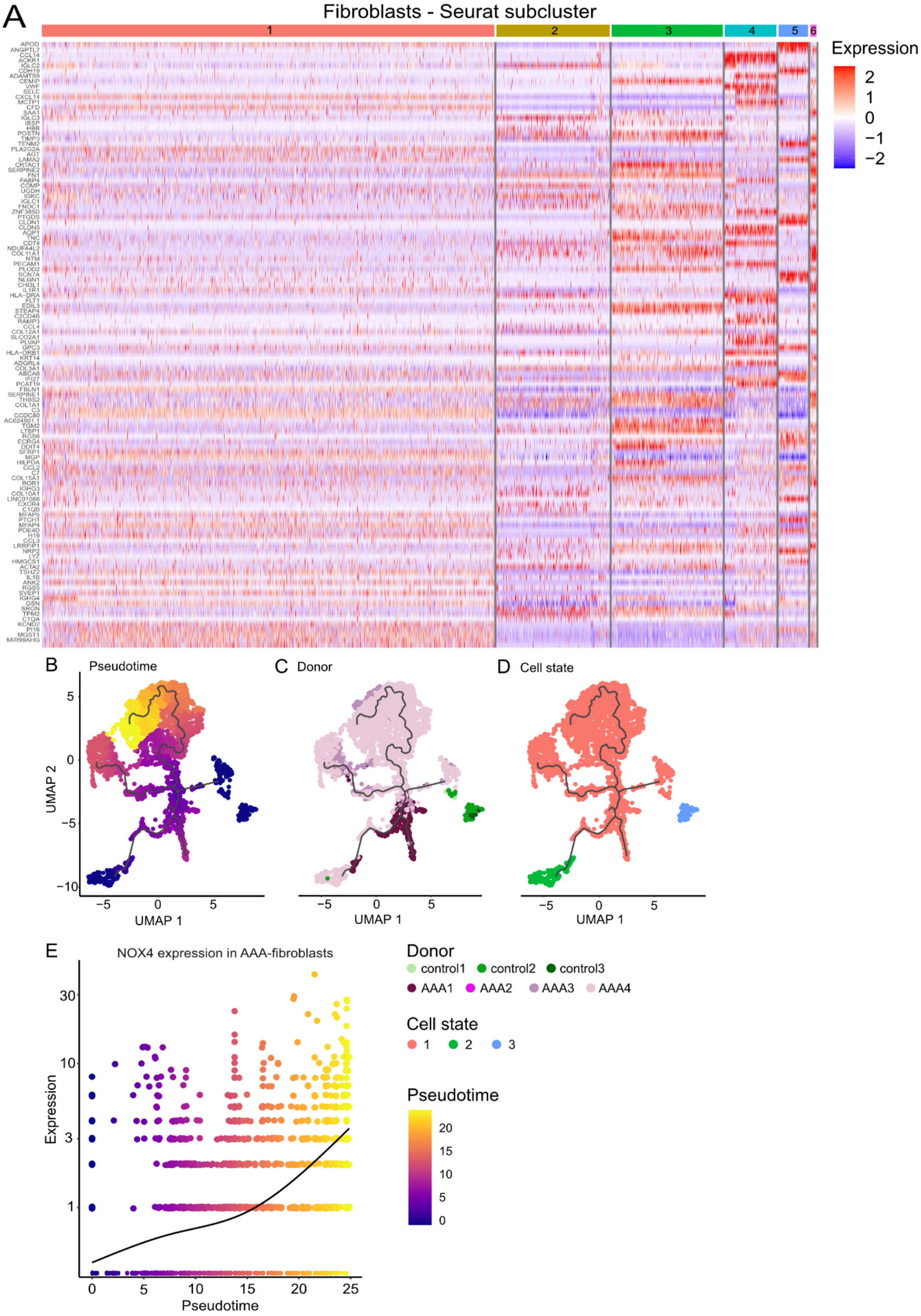
A
Fibroblasts - Seurat subcluster
1
2
3
4
5
6
Expression
2
1
0
−1
−2
B
Pseudotime
UMAP 2
UMAP 1
C
Donor
UMAP 1
D
Cell state
UMAP 1
E
NOX4 expression in AAA-fibroblasts
Expression
Pseudotime
Donor
control1 control2 control3
AAA1 AAA2 AAA3 AAA4
Cell state
1 2 3
Pseudotime
20
15
10
5
0

**Supplementary Figure 6: Characteristics of fibroblast Seurat subclusters and pseudotime trajectory of fibroblasts and NOX4 kinetics in human AAA and non-diseased controls. A,** Heatmap showing differentially expressed genes among the six fibroblast subpopulations in all integrated AAA ordered by log2 fold changes (scale). Colour scheme is based on relative gene expression (z-score) and comparison of the selected cluster with all others. Single cell trajectories are colored **B,** by pseudotime, **C,** by the different AAA patients (AAA1-AAA4) and non-diseased controls (control 1-control 3), and **D,** by the cell states (1-3). Black lines on the UMAP plots represent the trajectory graph. **E,** Pseudotime kinetics of NOX4 expression in fibroblasts from diseased AAA patients. Expression is expressed in log10. Pseudotime passes through a range of colours to represent progress through the transition. Dark blue = root cells, yellow = end cells. Electively treated abdominal aortic aneurysm (eAAA; n=4) were included in the analysis. All samples represent biological replicates.

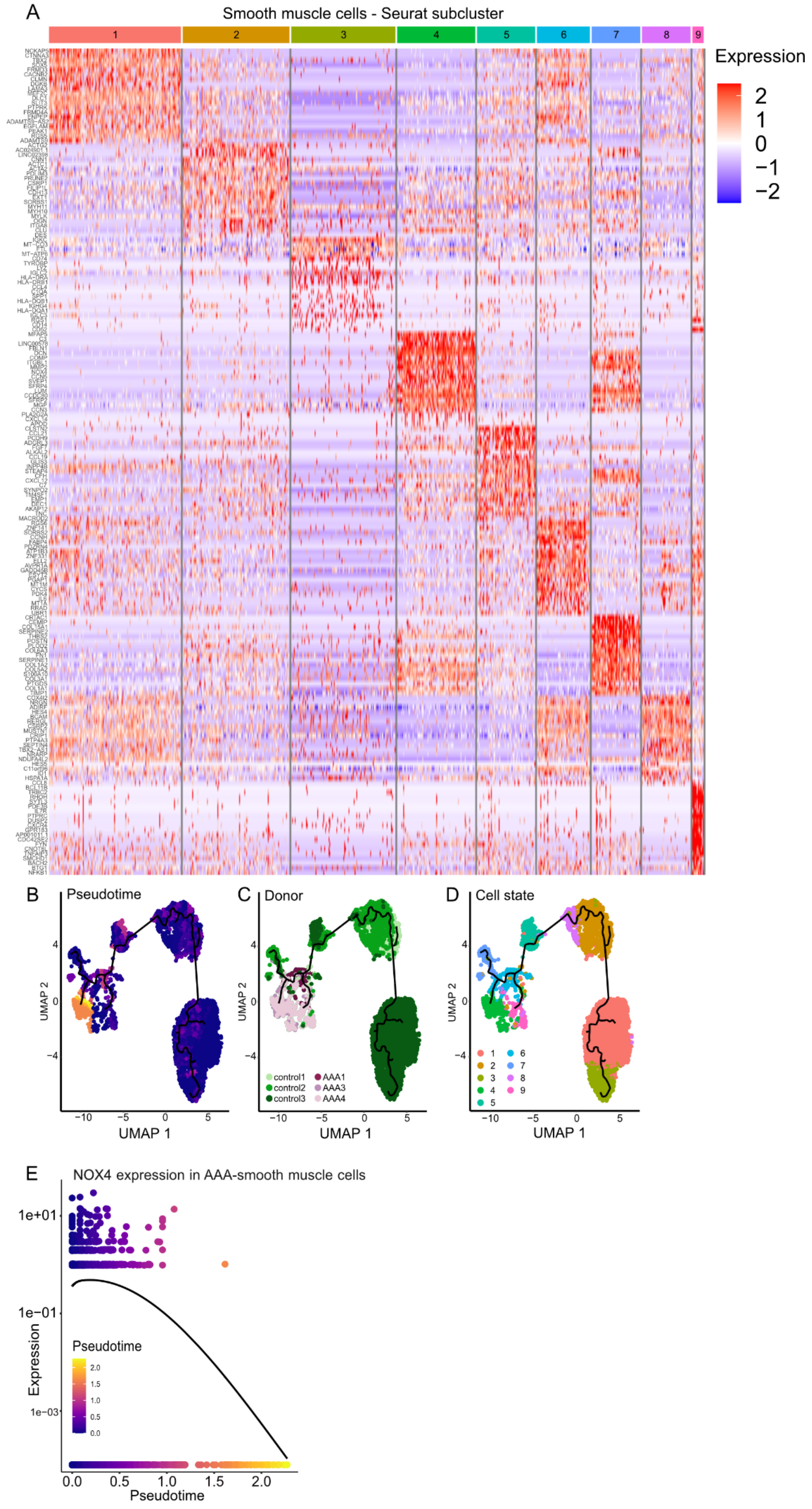
A
Smooth muscle cells - Seurat subcluster
1
2
3
4
5
6
7
8
9
Expression
2
1
0
−1
−2
B
Pseudotime
UMAP 2
UMAP 1
C
Donor
control1
control2
control3
AAA1
AAA3
AAA4
D
Cell state
E
NOX4 expression in AAA-smooth muscle cells
1e+01
1e−01
1e−03
Expression
Pseudotime
0.0
0.5
1.0
1.5
2.0

**Supplementary Figure 7: Characteristics of smooth muscle cell Seurat subclusters and pseudotime trajectory of smooth muscle cells and NOX4 kinetics in human AAA and non-diseased controls. A,** Heatmap showing differentially expressed genes among the six fibroblast subpopulations in all integrated AAA ordered by log2 fold changes (scale). Colour scheme is based on relative gene expression (z-score) and comparison of the selected cluster with all others. Single cell trajectories are colored **B,** by pseudotime, **C,** by the different AAA patients (AAA1-AAA4) and non-diseased controls (control 1-control 3), and **D,** by the cell states (1-3). Black lines on the UMAP plots represent the trajectory graph. **E,** Pseudotime kinetics of NOX4 expression in fibroblasts from diseased AAA patients. Expression is expressed in log10. Pseudotime passes through a range of colours to represent progress through the transition. Dark blue = root cells, yellow = end cells. Electively treated abdominal aortic aneurysm (eAAA; n=4) were included in the analysis. All samples represent biological replicates.

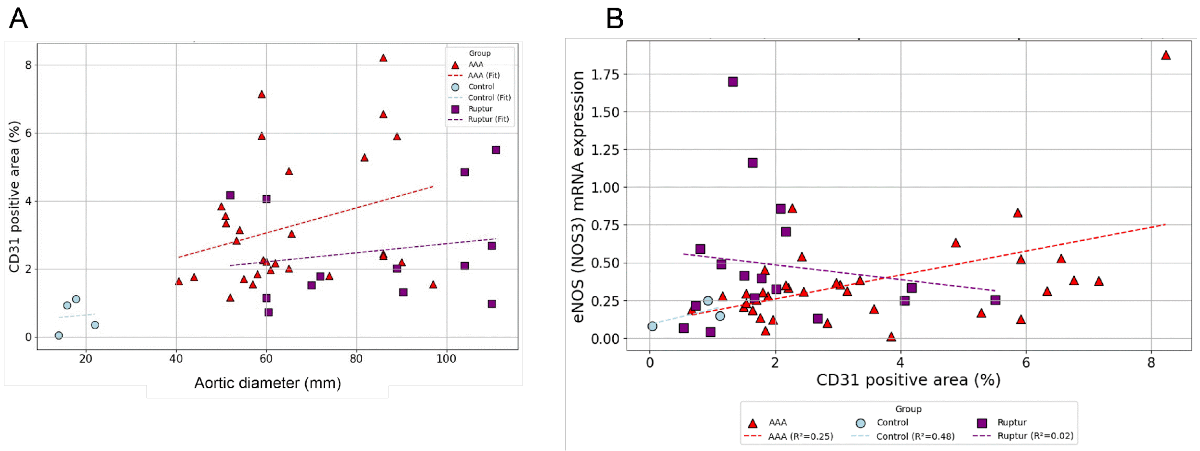


**Supplementary Figure 8: Correlation of CD31-positive vessels with the aortic diameter and eNOS expression in human AAA and corresponding controls.** Aortic specimens were obtained from patients undergoing elective surgical repair (eAAA, red dots), patients with ruptured AAA (rAAA, purple dots) and non-diseased controls (Ctrl, blue dots). **A,** CD31-positive vessels were quantified by immunohistochemistry. Correlation between aortic diameter and the number of CD31-positive vessels per mm² and aortic diameter. Controls (n=4), electively abdominal aortic aneurysm (eAAA; n=28), and ruptured AAA (rAAA; n=13) samples were included in the analysis. All samples represent biological replicates. **B,** Associations between the CD31-positive area with eNOS (NOS3) mRNA expression. Endothelial cells were stained with CD31 using immunohistochemistry and the CD31-positive area was quantified using a macro in Image J. eNOS mRNA expression was quantified by qPCR. Controls (n=3), electively abdominal aortic aneurysm (eAAA; n=35), and ruptured AAA (rAAA; n=17) samples were included in the analysis. All samples represent biological replicates.

A

Endothelial cells - Seurat subcluster

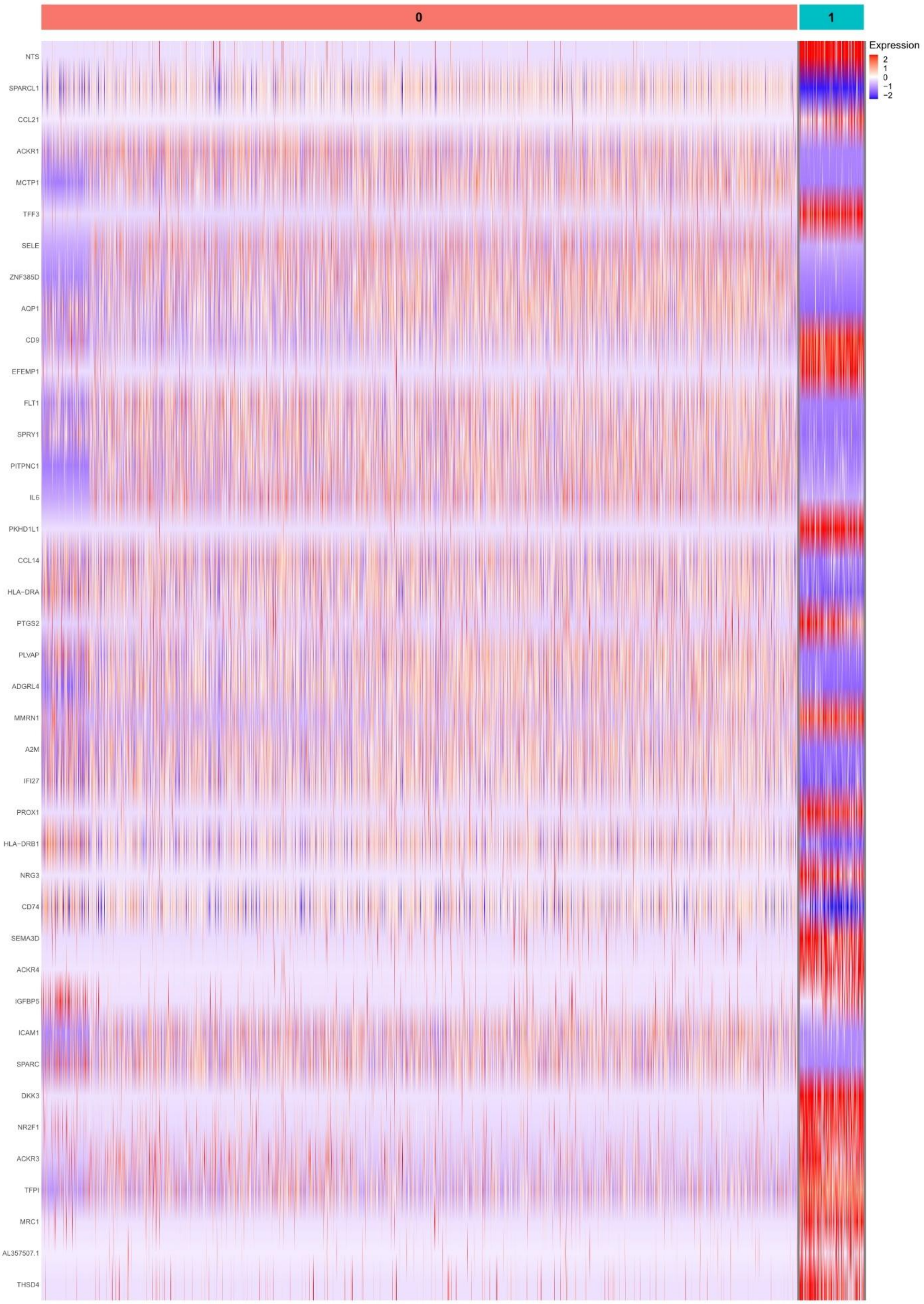

**Supplementary Figure 9: Characteristics of endothelial cell subclusters in human AAA**. Heatmap showing differentially expressed genes among the two endothelial cell subpopulations in all integrated AAA ordered by log2 fold changes (scale). Color scheme is based on relative gene expression (z-score) and comparison of the selected cluster vs all others. Electively treated abdominal aortic aneurysm (eAAA; n=4) were included in the analysis. All samples represent biological replicates.

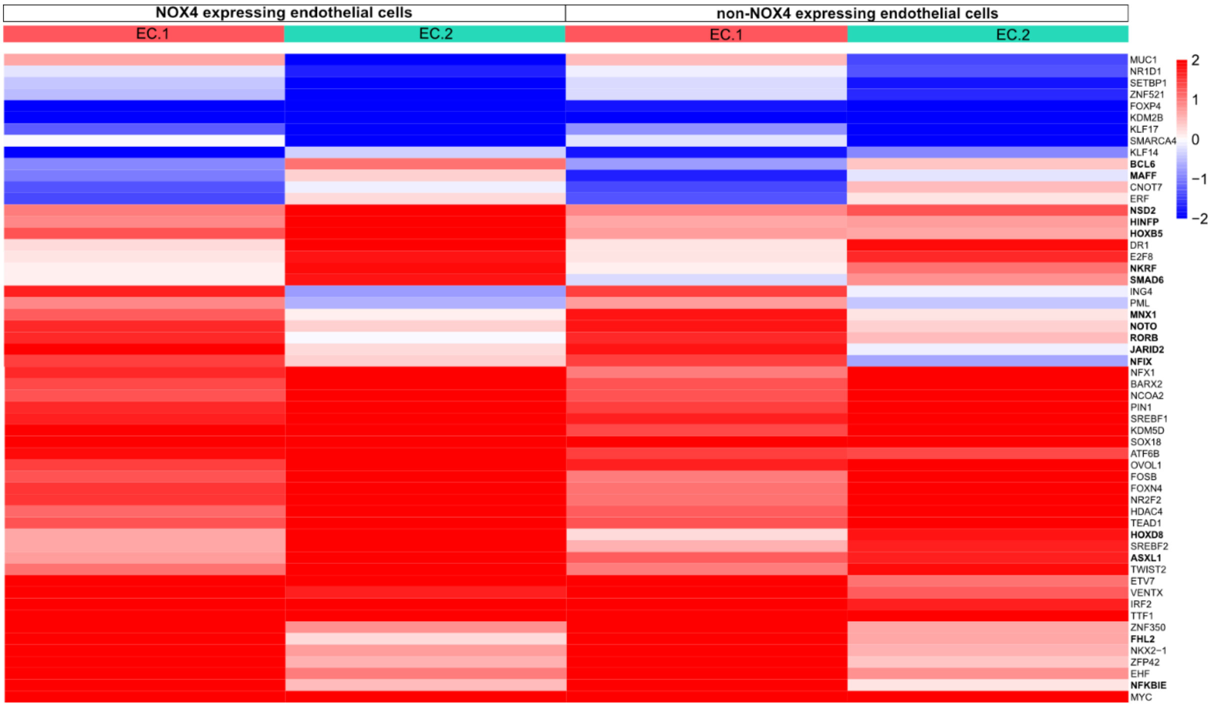


**Supplementary Figure 10: Prediction of transcription factor activities in Seurat subclusters of endothelial cells.** Heatmap visualizing the x-fold changes in activity of transcription factors between EC.1 and EC.2 expressing NOX4 (NOX4-expressing endothelial cells) or non-NOX4-expressing (non-NOX4-expressing endothelial cells). Each row presents a transcription factor and each column a cell with and without NOX4 expression in the different Seurat subclusters. The heatmaps were generated by aggregated scATAC-seq data as input and the predicted regulon activity and transcription factor gene expression are shown. Transcription factors are grouped based on specific cluster expression and cells are ordered based on clustering. Expression is log10 based. Red: upregulation, blue: downregulation. Electively treated abdominal aortic aneurysm (eAAA; n=4) were included in the analysis. All samples represent biological replicates.

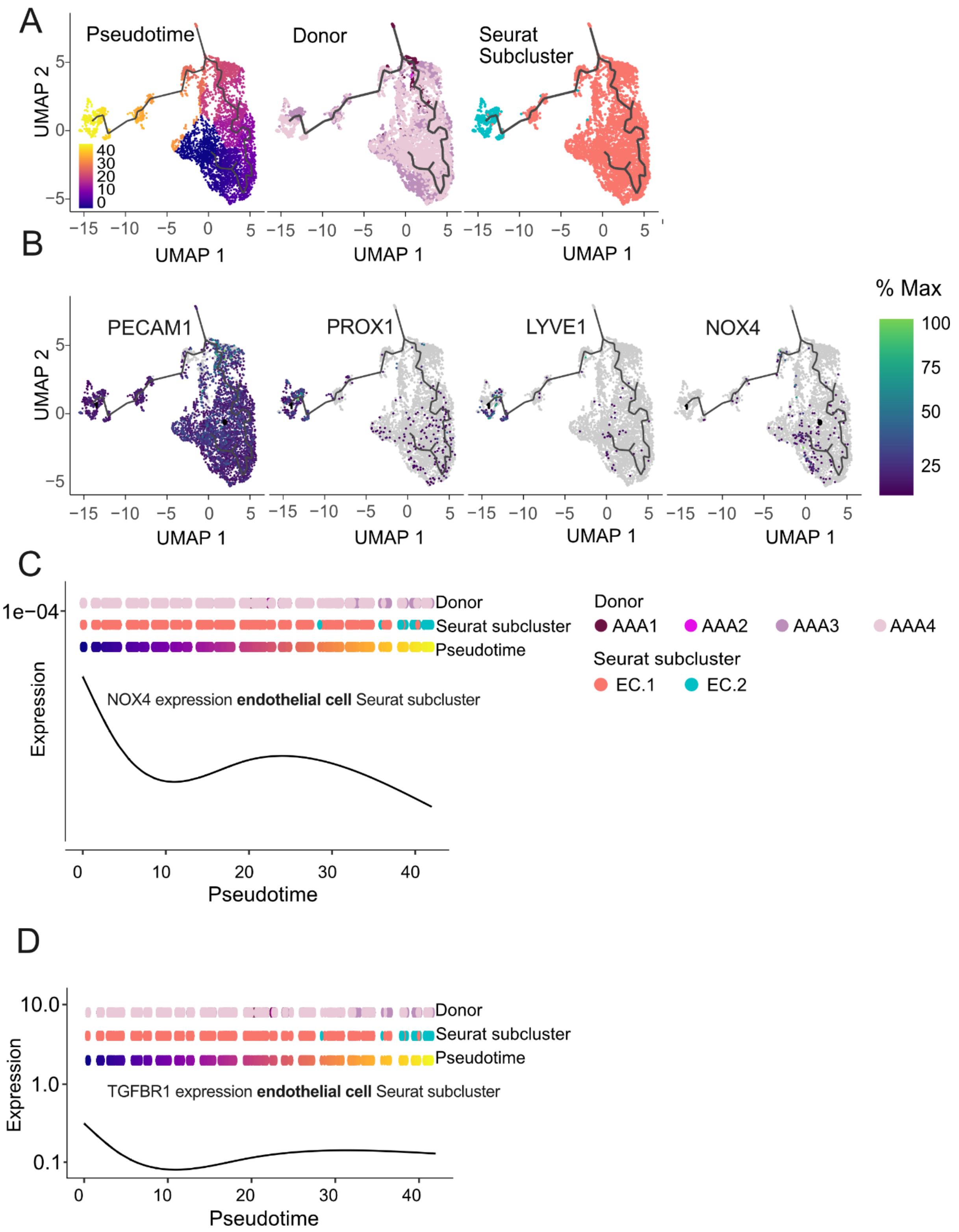


**Supplementary Figure 11: Pseudotime trajectory and NOX4 and TGFBR1 kinetics in Seurat subclusters of human AAA endothelial cell.** Single-cell trajectories coloured by **A,** pseudotime, by the different AAA donors (AAA.1-AAA.4) and by Seurat subclusters (EC.1-EC.1). Cells are plotted on a UMAP and the black lines on the UMAP plots represent the trajectory graph. **B,** Feature plots of selected genes in the different Seurat clusters during pseudotime. Pseudotime kinetics of **C,** NOX4 and **D,** TGFBR1 expression split by donor, endothelial cell Seurat subcluster and by pseudotime. Expression is based on log10. Pseudotime passes through a range of colours to represent progress through the transition. Dark blue = root cells, yellow = end cells. Electively treated abdominal aortic aneurysm (eAAA; n=4) were included in the analysis. All samples represent biological replicates.

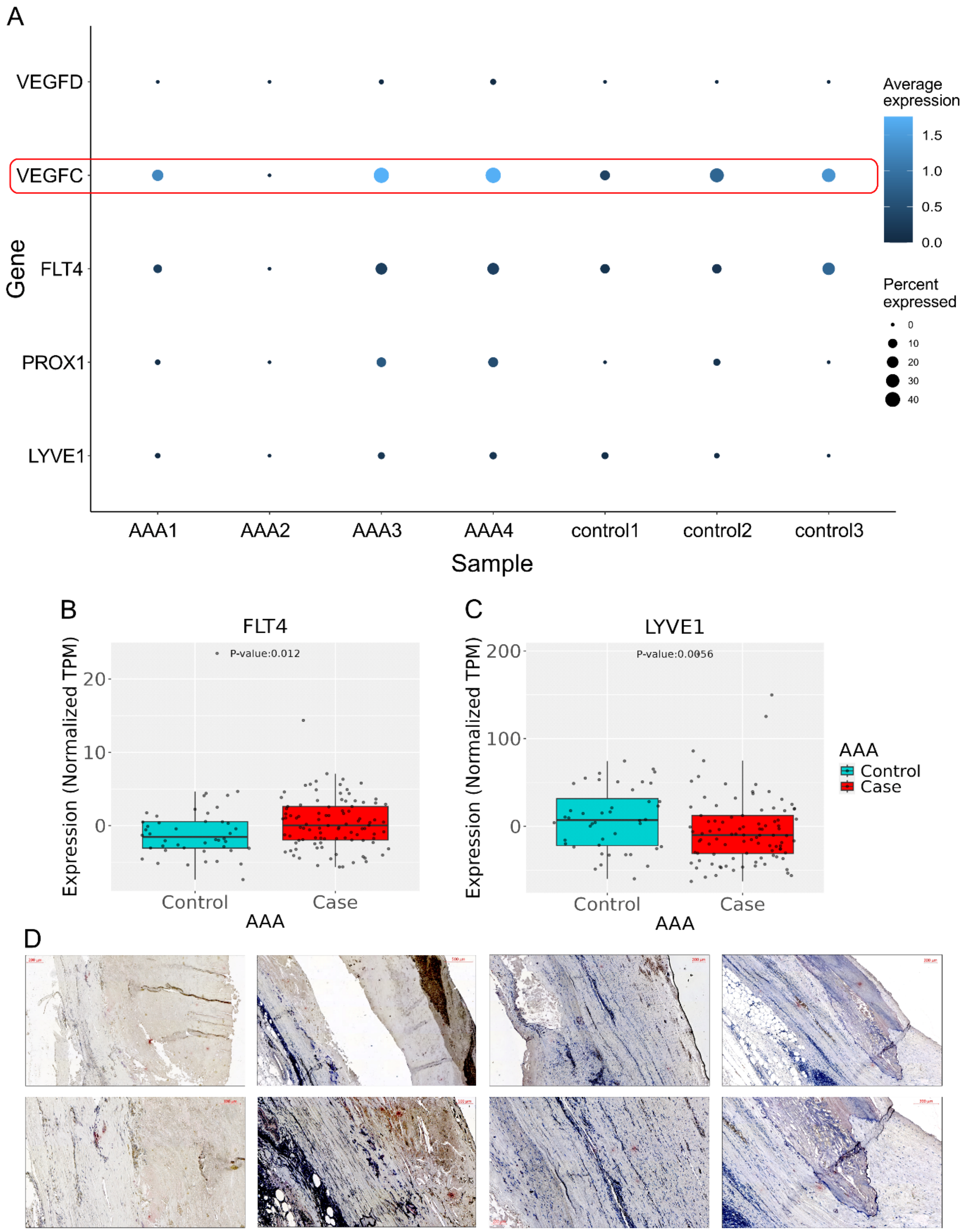


**Supplementary Figure 12: Differential gene expression of lymphangiogenic markers in AAA and non-AAA controls. A,** Feature plot of relative expression of LYVE1, PROX1, FLT4, VEGFC and VEGFD in endothelial cells in human AAA1-AAA4 and corresponding non-AAA controls (control1-control3). Electively treated abdominal aortic aneurysm (eAAA; n=4) were included in the analysis.

All samples represent biological replicates. Box plots comparing **B,** FLT4 and **C,** LYVE1 expression in AAA (case, n=96) and healthy controls (control, n=44) using data from bulk RNA-sequencing from whole aortic tissue. The horizontal line represents the median and the box represents the interquartile range. Expression is presented in transcripts per million (TPM) normalized for gene length and for sequencing depth. Results are adjusted for age, sex and technical covariates. **D,** Representative PROX1 immunohistochemistry in human AAA tissue (n=4, biological replicates). The red stars mark the DAB-positive areas. Red/brown = DAB-positive areas; blue = hemalum positive cell nuclei. Scale bars represent 100 µm, 200 µm, and 500 µm, as indicated in the figure.

#### Supplementary Tables

**Supplementary Table 1. Correlations of NOX4 mRNA with inflammation and histopathological vessel wall generation in ruptured AAA.** NOX4 and COL3A1 mRNA expression was analyzed by qPCR and the cleaved caspase-3 by immunohistochemistry. Elastin degradation was assessed semiquantitatively by six persons. The DNA binding activity of the transcription factor NF-κB was assessed by the TransAM NF-κB DNA-binding ELISA using nuclear extracts from AAA tissues. Electively treated abdominal aortic aneurysm (eAAA; n≥9) were included in the analysis. All samples represent biological replicates.

| | **NOX4 mRNA expression** |
|---|---|
| **Cleaved caspase-3, % positive area** | r= 0.7015<br>p= 0.0094<br>n= 13 |
| **Elastin degradation score** | r= 0.8398<br>p<0.0001<br>n= 19 |
| **COL3A1 mRNA expression** | r= 0.5516<br>p= 0.0438<br>n= 14 |
| **NF-κB binding activity** | r= 0.7280<br>p= 0.0313<br>n= 9 |

**Supplementary Table 2. Selected differential gene expression (DEG), GO and KEGG analysis of the DEG in the different fibroblasts subclusters.** ↑, gene sets the biological process are enriched; ↓gene sets the biological process are decreased. Electively treated abdominal aortic aneurysm (eAAA; n=4) were included in the analysis. All samples represent biological replicates.

| | DEG | Functional pathway analysis |
|---|---|---|
| Fibro.1 | ACTA2 low, CXCL14 | *Calcineurin-NFAT signaling cascade↑<br>*Cellular response to laminar fluid shear stress↑<br>*TGF-beta signaling pathway↑<br>*Activation Of AP-1 family of transcription factors↑<br>*Vascular Associated Smooth Muscle Cell Development↓<br>*Venous Blood Vessel Morphogenesis↓ |
| Fibro.2 | ACTA2, SRGN, TPM, TYH1 | *Cytoplasmic Translation↑<br>*Proton Motive Force-Driven Mitochondrial ATP Synthesis↑<br>*Regulation Of RNA Export From Nucleus↓<br>*Ribosome↑<br>*Adherens junction↓ |
| Fibro.3 | POSTN, FN1, PLOD, CRTAC1, EDIL3, STEAP4,<br>THBS2, CXCL14, C3, CFD, PLA2G2A, CDC80, MGP | *Positive regulation of vascular endothelial growth factor signaling pathway↑<br>*Positive regulation of cardiac epithelial to mesenchymal transition↑<br>*Focal adhesion↑<br>*Crosslinking of collagen fibril↑<br>*RUNX3 regulates WNT signaling↑ |

| | | |
|---|---|---|
| | | *RUNX2 regulates genes involved in cell migration↑<br>*Cytoplasmic translation↓ |
| Fibro.4 | CCL14, ACKR1, VWF, CLND5, AQP1, PECAM1, MYL9, TAGLN, ACTA2, HLA-DRA, HLA-DRB1, CD74 | *Cytoplasmic Translation↑<br>*Ribosome↑<br>*Various types of N-glycan biosynthesis↓ |
| Fibro.5 | APOD, TENM2, CLDN1, SCN7A, NRP2, CPC3, CDH19, LAMA2 | *Regulation Of Basement Membrane Organization↑<br>*Cholesterol Biosynthesis ↑<br>*ECM-receptor interaction↓<br>*Crosslinking Of Collagen Fibrils↓ |
| Fibro.6 | TIMP3, TIMP1, SERPINE1, SERPINE2, NTM, NDUFA4L2, UGDH | *Negative regulation of plasminogen activation↑<br>*Embryonic eye morphogenesis↑<br>*Protein digestion and absorption↑<br>*Collagen chain trimerization↑<br>*Cell cycle↓ |

**Supplementary Table 3. Top 5 up-regulated GO enrichment analysis in Seurat subcluster - Fibroblasts.** Functional analysis by GO (Top5 GO terms) for upregulated differentially expressed genes in the indicated fibroblast subcluster vs. the other fibroblasts subclusters. Electively treated abdominal aortic aneurysm (eAAA; n=4) were included in the analysis. All samples represent biological replicates.

| **Fibro.1** | |
|---|---|
| Calcineurin-NFAT Signaling Cascade | GO:0033173 |

| | |
|---|---|
| Cellular Response To Laminar Fluid Shear Stress | GO:0071499 |
| Response To Heparin | GO:0071503 |
| Regulation Of Keratinocyte Apoptotic Process | GO:1902172 |
| Blood Vessel Endothelial Cell Proliferation Involved In Sprouting Angiogenesis | GO:0002043 |
| **Fibro.2** | |
| Cytoplasmic Translation | GO:0002181 |
| Proton Motive Force-Driven Mitochondrial ATP Synthesis | GO:0042776 |
| Proton Motive Force-Driven ATP Synthesis | GO:0015986 |
| Mitochondrial Electron Transport, NADH To Ubiquinone | GO:0006120 |
| Protein Insertion Into ER Membrane By Stop-Transfer Membrane-Anchor Sequence | GO:0045050 |
| **Fibro.3** | |
| Positive Regulation Of Vascular Endothelial Growth Factor Signaling Pathway | GO:1900748 |
| Positive Regulation Of Cardiac Epithelial To Mesenchymal Transition | GO:0062043 |
| Peptidyl-Lysine Hydroxylation | GO:0017185 |
| Regulation Of Endoplasmic Reticulum Tubular Network Organization | GO:1903371 |
| Negative Regulation Of Cellular Response To Hypoxia | GO:1900038 |

| | |
|---|---|
| **Fibro.4** | |
| Cytoplasmic Translation | GO:0002181 |
| Endothelial Cell Development | GO:0001885 |
| Vascular Associated Smooth Muscle Cell Development | GO:0097084 |
| Regulation Of Cell Adhesion Molecule Production | GO:0060353 |
| Positive Regulation Of CD4-positive, CD25-positive, Alpha-Beta Regulatory T Cell Differentiation | GO:0032831 |
| **Fibro.5** | |
| Regulation Of Basement Membrane Organization | GO:0110011 |
| Positive Regulation Of Tau-Protein Kinase Activity | GO:1902949 |
| Positive Regulation Of Sterol Biosynthetic Process | GO:0106120 |
| Positive Regulation Of Cholesterol Biosynthetic Process | GO:0045542 |
| Negative Regulation Of Erythrocyte Differentiation | GO:0045647 |
| **Fibro.6** | |
| Negative Regulation Of Plasminogen Activation | GO:0010757 |
| Embryonic Eye Morphogenesis | GO:0048048 |
| Positive Regulation Of Extrinsic Apoptotic Signaling Pathway Via Death Domain Receptors | GO:1902043 |
| Receptor-Mediated Virion Attachment To Host Cell | GO:0046813 |

| | |
|---|---|
| Negative Regulation Of Membrane Protein Ectodomain Proteolysis | GO:0051045 |

**Supplementary Table 4. Top 5 down-regulated GO enrichment analysis in Seurat subcluster - Fibroblasts.** Functional analysis by GO (Top5 GO terms) for downregulated differentially expressed genes in the indicated fibroblast subcluster vs. the other fibroblasts subclusters. Electively treated abdominal aortic aneurysm (eAAA; n=4) were included in the analysis. All samples represent biological replicates.

| | |
|---|---|
| **Fibro.1** | |
| Mitochondrial Electron Transport, Cytochrome C To Oxygen | GO:0006123 |
| Negative Regulation Of Trophoblast Cell Migration | GO:1901164 |
| Vascular Associated Smooth Muscle Cell Development | GO:0097084 |
| Calcitonin Family Receptor Signaling Pathway | GO:0097646 |
| Venous Blood Vessel Morphogenesis | GO:0048845 |
| **Fibro.2** | |
| Regulation Of RNA Export From Nucleus | GO:0046831 |
| Calcineurin-NFAT Signaling Cascade | GO:0033173 |
| Regulation Of Ribonucleoprotein Complex Localization | GO:2000197 |
| Regulation Of mRNA Export From Nucleus | GO:0010793 |
| Regulation Of Basement Membrane Organization | GO:0110011 |
| **Fibro.3** | |
| Cytoplasmic Translation | GO:0002181 |

| | |
|---|---|
| Peptide Biosynthetic Process | GO:0043043 |
| Positive Regulation Of Intrinsic Apoptotic Signaling Pathway By P53 Class Mediator | GO:1902255 |
| Response To Heparin | GO:0071503 |
| Regulation Of Type B Pancreatic Cell Proliferation | GO:0061469 |
| **Fibro.4** | |
| Peptidyl-Proline Hydroxylation To 4-hydroxy-L-proline | GO:0018401 |
| Golgi Transport Vesicle Coating | GO:0048200 |
| COPI-coated Vesicle Budding | GO:0035964 |
| COPI Coating Of Golgi Vesicle | GO:0048205 |
| Negative Regulation Of Dendritic Spine Development | GO:0061000 |
| **Fibro.5** | |
| Regulation Of Epithelial To Mesenchymal Transition Involved In Endocardial Cushion Formation | GO:1905005 |
| Receptor-Mediated Virion Attachment To Host Cell | GO:0046813 |
| Retinal Ganglion Cell Axon Guidance | GO:0031290 |
| Glycolipid Transport | GO:0046836 |
| B Cell Chemotaxis | GO:0035754 |
| **Fibro.6** | |

| | |
|---|---|
| Positive Regulation Of Establishment Of Protein Localization To Telomere | GO:1904851 |
| TORC2 Signaling | GO:0038203 |
| Regulation Of Protein Targeting | GO:1903533 |
| Negative Regulation Of Stress-Activated Protein Kinase Signaling Cascade | GO:0070303 |
| Regulation Of Protein Localization To Cilium | GO:1903564 |

**Supplementary Table 5. Regulated KEGG pathways in Seurat subclusters – Fibroblasts.** Top5 significantly enriched KEGG pathways for upregulated genes and downregulated genes in the indicated fibroblast subcluster vs. the other fibroblasts subclusters. Electively treated abdominal aortic aneurysm (eAAA; n=4) were included in the analysis. All samples represent biological replicates.

| **Fibro.1** | |
|---|---|
| **Up-Regulated** | **Down-Regulated** |
| AGE−RAGE signaling pathway in diabetic complications | Oxidative phosphorylation |
| TGF−beta signaling pathway | Proteasome |
| Fatty acid degradation | Viral myocarditis |
| Circadian rhythm | Type I diabetes mellitus |
| Nicotinate and nicotinamide metabolism | Allograft rejection |
| **Fibro.2** | |
| Ribosome | Adherens junction |
| Parkinson disease | Hedgehog signaling pathway |
| Oxidative phosphorylation | Renal cell carcinoma |
| Proteasome | Circadian rhythm |
| Protein export | Phosphonate and phosphinate metabolism |
| **Fibro.3** | |

| | |
|---|---|
| Focal adhesion | Ribosome |
| Bacterial invasion of epithelial cells | Coronavirus disease |
| Renal cell carcinoma | Fatty acid degradation |
| Neomycin, kanamycin and gentamicin biosynthesis | Sulfur metabolism |
| D-Glutamine and D-glutamate metabolism | Phosphonate and phosphinate metabolism |
| **Fibro.4** | |
| Ribosome | Various types of N-glycan biosynthesis |
| Viral myocarditis | N-Glycan biosynthesis |
| Allograft rejection | Protein export |
| Graft-versus-host disease | Glycosaminoglycan degradation |
| Type I diabetes mellitus | Vitamin B6 metabolism |
| **Fibro.5** | |
| Parathyroid hormone synthesis, secretion and action | Viral myocarditis |
| Glioma | ECM-receptor interaction |
| ErbB signaling pathway | Hypertrophic cardiomyopathy |

| | |
|---|---|
| Terpenoid backbone biosynthesis | AGE−RAGE signaling pathway in diabetic complications |
| Steroid biosynthesis | Arrhythmogenic right ventricular cardiomyopathy |
| **Fibro.6** | |
| Protein digestion and absorption | Proteasome |
| ECM−receptor interaction | Cell cycle |
| AGE−RAGE signaling pathway in diabetic complic | Aldosterone−regulated sodium reabsorption |
| Amoebiasis | Sulfur metabolism |
| Small cell lung cancer | Terpenoid backbone biosynthesis |

**Supplementary Table 6. Selected differential gene expression (DEG), GO and KEGG analysis of the DEG in the different smooth muscle cell subclusters.** ↑, gene sets the biological process are enriched; ↓gene sets the biological process are decreased. Electively treated abdominal aortic aneurysm (eAAA; n=4) were included in the analysis. All samples represent biological replicates.

| | DEG | Functional pathway analysis |
|---|---|---|
| SMC.1 | RGS5, SOX5, | *Elastic Fiber Assembly↑<br>*ECM receptor interaction↑ |
| SMC.2 | MYH11, ACTA2, CNN1, OGN, CDH13 | *Elastic Fiber Assembly↑ |
| SMC.3 | CD74, IGLC2, HLA-DRA, HLA-DRB1, IGLC3 | *Positive Regulation Of Extracellular Matrix Assembly↑ |
| SMC.4 | FBLN1, DCN, LUM, CCDC80, MGP, C3, COMP, MMP2, MYLK, FBLN1, MFAP5, NT5E, ENG, THY1 | *Elastic fiber assembly↑<br>*Regulation of autophagy↑<br>*Positive regulation of cardiac muscle cell differentiation↓<br>*Vascular associated smooth muscle cell development↓<br>*Vascular smooth muscle contraction↑<br>*Glycosaminoglycan degradation↓ |
| SMC.5 | CLSTN2, CCL21, PCDH9, CCL19, C7, DEC1 | *Negative Regulation Of Vascular Permeability↑<br>*Cytoplasmic Translation↓<br>*Ribosome↑ |
| SMC.6 | RGS6, CCNH, FABP4, ZNF141, ZNF331, IL6, DGKB | *Regulation Of Mesenchymal Stem Cell Differentiation<br>*Vascular Associated Smooth Muscle Cell Differentiation |
| SMC.7 | CRTAC1, POSTN, CEMIP, NT5E, ENG, THY1 | *Protein Retention in ER Lumen↑<br>*Cytoplasmic Translation↓ |

| | | |
|---|---|---|
| | | *Protein processing in endoplasmic reticulum↓ |
| SMC.8 | NRGN, HES4, SEPTIN4, | *Cytoplasmic Translation↑<br>*Histone Lysine Demethylation ↓<br>*Adherens junction↑ |
| SMC.9 | RGS1, CD14, BCL11B, RHOH, IL7R, CXCR4, NFKB1, BTG1, BACH2, DUSP2 | *Histone Lysine Demethylation↑<br>*Interleukin-2-Mediated Signaling Pathway↑<br>*Cellular Response To Interleukin-2↑<br>*Vascular smooth muscle contraction↑<br>*Th1 and Th2 cell differentiation↓<br>*Th17 cell differentiation↓ |

**Supplementary Table 7. Selected differential gene expression (DEG) and GO and KEGG analysis of the DEG in the different endothelial cell subclusters.** ↑, gene sets the biological process are enriched; ↓gene sets the biological process are decreased. Electively treated abdominal aortic aneurysm (eAAA; n=4) were included in the analysis. All samples represent biological replicates.

| | **DEG** | **Functional pathway analysis** |
|---|---|---|
| EC.1 | SELE, ICAM1, HLA-DRA, SPARC, MCTP1, ACKR1, ZNF385D, CCL14, SPRY1 | *Interleukin-15-Mediated Signaling Pathway↑<br>*Cellular Response to Interleukin-15↑<br>*TH17 cell differentiation↑<br>*Interleukin-15 Signaling↑<br>*Interleukin-6 Signaling↑ |
| EC.2 | LYVE1, PROX1, TFF3, EFEMP1, CD9, MMRN1, DKK3, NR2F1, ACKR3, MRC1, THSD4 | *Proton Motive Force-Driven Mitochondrial ATP Synthesis↑<br>*Oxidative Phosphorylation↑ |

| | | |
|---|---|---|
| | | *Formation of ATP by Chemiosmotic Coupling↑<br>*Cholesterol Biosynthesis↑ |

**Supplementary Table 8. Selected transcription factor activity in the different identified cell clusters in patients with AAA.** Electively treated abdominal aortic aneurysm (eAAA; n=4) were included in the analysis. All samples represent biological replicates.

| | **TF increased activity** | **TF reduced activity** |
|---|---|---|
| **NOX4-expressing cells vs. non-NOX4-expressing cells** | SMAD3, SRF, TP53, GATA4, CTNNB1, MRTFA, HOXA5, FOSL1, FEZF2, TWIST1, TFAP2A | TCF7L1, SMAD7, HDAC5, ZEB1 |
| **NOX4-expressing cells vs. non-NOX4-expressing smooth muscle cells** | ATF1, ATF3, ATF4, ATF5, CEBPA, CEBPB, CEBPE, CEBPG, CTNNB1, DLX1, ELF4, ETS1, ETS2, FOS, FOSL1, FOXF1, FOXO1, FOXO3, GATA4, HIF1A, HOXA5, HOXD3, HOXD13, IRF2, JUNB, KLF5, MEF2C, MITF, MYB, MYOD1, NCOR1, NFAT5, NFATC1, NFE2L2, NFKB1, NR3C2, NR4A1, RXRB, SMAD2, SMAD3, SMAD4, SNAI1, SOX18, SP1, SP3, SPI1, SRF, STAT1, STAT3, TFAP2A, TLX3, TP53, TWIST1, ZEB1 | NR1D1, SNAI2 |
| **NOX4-expressing cells vs. non-NOX4-expressing fibroblasts** | ATF1, ATF2, ATF3, ATF4, ATF5, ATF6, ATF6B, ATOH8, CEBPA, CEBPB, CEBPD, CEBPE, CEBPG, CTNNB1, DLX1, E2F1, EGR2, ELF1, ELF3, ELF4, ELF5, EP300, ETS1, ETS2, FOS, FOSB, FOSL1, FOXA3, | IRF5, IRF7, LMO2, NFE2L3, NR1D1, SMAD7, SMARCA4, SNAI2, SOX18 |

| | | |
|---|---|---|
| | FOXF1, FOXO3, GATA4, GLI1, HEY1, HIF1A, HIF3A, HOXA5, HOXD3, HOXD13, IRF1, IRF2, IRF3, IRF8, IRF9, IRX3, IRX4, JUN, JUNB, JUND, KLF4, KLF5, KLF11, MEF2C, MYB, MYOD1, MYOG, NANOG, NCOR1, NEUROD4, NFAT5, NFATC1, NFATC4, NFE2L2, NFKB1, NR3C2, NR4A1, PPARA, PPARD, PPARG, PRRX1, PRRX2, RUNX1, RXRB, SKI, SMAD2, SMAD3, SMAD4, SMAD6, SMARCA2, SNAI1, SOX10, SP1, SP3, SPI1, SRF, STAT1, STAT3, TBX4, TEAD1, TEAD2, TEAD3, TEAD4, TFAP2A, TLX3, TOX3, TP53, TWIST1, TWIST2, ZEB1, ZKSCAN7 | |
| **NOX4-expressing cells vs. non-NOX4-expressing endothelial cells** | ATF1, ATF2, ATF3, ATF4, ATF5, ATF6B, BMP2, CEBPB, CEBPE, CEBPG, CEBPZ, E2F1, TLX3, NFE2L2, E2F1, ELF1, ELF2, ELF4, EP300, ETS1, ETS2, FOS, FOXO3, HEY1, HIF1A, HIF3A, HOXA5, HOXD3, IRF3, IRF5, IRF8, JDP2, JUN, KLF5, KLF11, LMO2, MEF2C, MITF, MYB, MYOD1, NANOG, NCOR1, NEUROD4, NFAT5, | ATOH8, ATF7, DLX1, DLX5, FOSL1, IRF5, IRF6, IRF7, IRX3, IRX4, JUND, LHX9, NFE2L3, NFKB2, PPARG, PRRX1 |

| | | |
|---|---|---|
| | NFATC4, NFE2L2, NR3C2, NR4A1, RUNX1, SKI, SMAD2, SMAD3, SMAD6, SMARCA2, SMARCA4, SNAI1, SP1, SP3, SPI1, STAT1, STAT3, TAZ, TEAD4, TFAP2A, TGIF2, TLX3, TOX3, TP53, TWIST1, ZKSCAN7 | |
| **NOX4-expressing SMC.4 vs. non-NOX4-expressing SMC.4** | KLF8, NOTCH1 | |
| **NOX4-expressing SMC.7 vs. non-NOX4-expressing SMC.7** | KLF8, IRF3 | HDAC9 |
| **NOX4-expressing Fibro.3 vs. non-NOX4-expressing Fibro. 3** | MUC1 | |
| **NOX4-expressing Fibro.5 vs. non-NOX4-expressing Fibro.5** | SMAD7 | |
| **NOX4-expressing Fibro.4 vs. non-NOX4-expressing Fibro.4** | | NFE2L3 |
| **NOX4-expressing EC.1 vs. non-NOX4-expressing EC.1** | HOXB5, SMAD6, ING4, MAFF, MUC1, MAFF, PIN1, ATF6B, FOSB, FOXN4, NR2F2, HOXD8, TWIST2 | KLF17, MNX1, JARID2, NOTO, SETBP1, ZNF521, KLF17, BCL6, ERF, BARX2, KDM5D, OVOL1, HDAC4, ASXL1 |
| **NOX4-expressing EC.2 vs. other non-NOX4-expressing EC.2** | KLF14, BCL6, MAFF, NSD2, HINFP, HOXB5, E2F8, NKRF, SMAD6, JARID2, NFIX, ETV7, VENTX, NFKBIE | MUC1, NR1D1, ZNF521, CNOT7, ING4, PML, MNX1, RORB, FHL2 |

**Supplementary Table 9.** Aortic tissues were obtained from patients undergoing elective open abdominal aortic aneurysm repair (eAAA) or surgery due to ruptured AAA (rAAA) or aortic-occlusive disease (Crtl) where the aortic segment was obtained from the atherosclerosis-free insertion site of the femoral or bifemoral bypass. For some patients, data on medical therapies were not available at the time of tissue collection and the number of analyzed patients (n) varies. **Statistics:** Depending on normally testing, the data are presented as median with range or mean with standard deviation, as indicated in the table. The comparison of continuous variables in Ctrl, eAAA and rAAA was done using the Kruskal-Wallis and Dunn's multiple comparisons test or One-Way ANOVA and Holm-Šídák's multiple comparisons test when the data were normally distributed. The data for HDL-, LDL and total cholesterol, and non-fasting blood glucose were compared using the Mann Whitney test. **$P$<0.001, ***$P$<0.0001 eAAA/rAAA vs. Ctrl; ##$P$<0.001, #$P$<0.0001 rAAA *vs.* eAAA. Differences in the distribution of d (cardiovascular risk factors, medical therapies) across the three independent groups were compared by Fisher's exact test using a contingency table. **Abbreviations:** ACE indicates angiotensin-converting enzyme; ARB, angiotensin receptor blockers; ASA, acetylsalicylic acid; BMI, body mass index; CCB, calcium-channel blockers; CAD, coronary artery disease; CRP, C-reactive protein; HDL, high-density lipoprotein; LDL, low-density lipoprotein; PAD, peripheral artery disease; and T2D, type 2 diabetes. Controls (n=6), electively abdominal aortic aneurysm (eAAA; n=49), and ruptured AAA (rAAA; n=21) samples were included in the analysis. All samples represent biological replicates.

| | **Ctrl** | **elective AAA** | **ruptured AAA** | **P-value** |
|---|---|---|---|---|
| Age, mean±SD, years, n | 55.5±7.5<br>(6) | 65.3±7.9<br>(49)** | 74.5±6.8<br>(21)***# | |
| Sex, m:f, % male | 4:2, 67 | 43:6, 88 | 18:3, 86 | $P$=0.37 |
| AAA diameter, mean±SD, mm, n | 19.98±4.81<br>(6) | 62.56±14.03<br>(49)**** | 80.88±20.58<br>(17)****### | |
| BMI, median with | 23.30 | 27.40 | 24.75 | |

| | | | | |
|---|---|---|---|---|
| range, kg/m², n | (19.00-37.20)<br>(6) | (19.80-42.90)<br>49 | (20.80-40.00)<br>12 | |
| Smoking history<br>yes:no, % smoker | 5:1, 83 | 33:16, 67 | 5:10, 33 | *P*=0.03 |
| Hypertension,<br>yes:no, % | 4:2, 67 | 43:6, 87 | 13:2, 87 | *P*=0.37 |
| CAD, yes:no, % | 3:3, 50 | 17:32, 37 | 5:10, 33 | *P*=0.84 |
| PAD, yes:no, % | 6:0, 100 | 12:37, 25 | 3:12, 20 | *P*=0.0009 |
| Type 2 diabetes,<br>yes:no, % | 1:5, 17 | 9:40, 18 | 3:12, 20 | *P*>0.99 |
| CRP, median with<br>range, mg/dL,<br>n | 4.05<br>(1.20-18.0)<br>6 | 2.90<br>(0.50-127.8)<br>49 | 10.55<br>(1.30-269.4)##<br>18 | |
| TG, median with<br>range, mmol/L,<br>n | 1.57<br>(0.79-1.79)<br>5 | 1.59<br>(0.73-6.69)<br>41 | 1.70<br>(1.44-2.29)<br>3 | ns |
| HDL-C, mmol/L,<br>n | 1.23 (0.85-1.48)<br>5 | 1.16 (0.65-2.43)<br>41 | – | |
| LDL-C, mmol/L,<br>n | 1.56 (1.26-5.08)<br>4 | 2.62 (0.71-6.79)<br>38 | – | ns |
| TC, mmol/L,<br>n | 3.02 (2.88-4.19)<br>4 | 4.47 (2.05-8.07)<br>38 | – | ns |
| non-fasting glucose,<br>mmol/L,<br>n | 5.50 (4.40-<br>30.60)<br>5 | 5.42 (3.95-<br>10.68)<br>42 | – | ns |
| Statin, yes:no, % | 5:1, 83 | 33:16, 67 | 9:5, 64 | *P*=0.83 |
| ARB, yes:no, % | 2:4, 33 | 16:32, 33 | 4:10, 29 | *P*>0.99 |
| ACE, yes:no, % | 0:6, 0 | 23:26, 47 | 6:8, 43 | *P*=0.09 |

| | | | | |
|---|---|---|---|---|
| β-blocker, yes:no, % | 2:4, 33 | 22:27, 45 | 8:6, 57 | *P*=0.63 |
| CCB, yes:no, % | 0:6, 0 | 20:29, 41 | 3:11, 21 | *P*=0.07 |
| Therapeutic anticoagulation, yes:no, % | 4:2, 50 | 43:6, 20 | 13:2, 50 | *P*=0.04 |
| Diuretics, yes:no, % | 0:6, 0 | 18:31, 37 | 5:9, 36 | *P*=0.24 |
| ASA, yes:no, % | 6:0, 100 | 30:19, 61 | 9:5, 64 | *P*=0.21 |
| Insulin, yes:no, % | 1:5, 17 | 3:46, 6 | 0:14, 0 | *P*=0.46 |
| Type 2 diabetes treatment, yes:no, % | 1:5, 17 | 6:43, 12 | 1:13, 7 | *P*=0.70 |

**Supplementary Table 10. Clinical characteristics of patients with AAA for analysis of single-cell RNA sequencing.** Aortic tissues were obtained from patients undergoing elective open abdominal aortic aneurysm repair. **Abbreviations:** ACE indicates angiotensin-converting enzyme; ARB, angiotensin receptor blockers; ASA, acetylsalicylic acid; BMI, body mass index; CCB, calcium-channel blockers; CAD, coronary artery disease; CRP, C-reactive protein; HDL, high-density lipoprotein; LDL, low-density lipoprotein; PAD, peripheral artery disease; and T2D, type 2 diabetes. Electively treated abdominal aortic aneurysm (eAAA; n=4) were included in the analysis. All samples represent biological replicates.

| | **AAA1** | **AAA2** | **AAA3** | **AAA4** |
|---|---|---|---|---|
| Age, years | 66-71 | | | |
| Sex | male | | | |
| AAA diameter, mm | 57.0 | 61.0 | 59.0 | 66.6 |
| BMI, $kg/m^2$ | 24.9 | 24.5 | 30.7 | 24.8 |
| Smoking history | yes | yes | no | yes |
| Hypertension | yes | yes | yes | yes |
| Alcohol abuse | no | yes | no | no |
| CAD | no | no | no | no |
| PAD | no | no | no | yes |
| Carotid artery stenosis | no | no | no | no |
| Type 2 diabetes | no | no | no | no |
| CRP, mg/dL | 1.8 | 0.5 | 3.5 | 0.5 |
| TG, mmol/L | 2.25 | 1.32 | 0.99 | 1.32 |
| HDL-C, mmol/L | 1.41 | 1.64 | 0.87 | 1.10 |

| | | | | |
|---|---|---|---|---|
| LDL-C, mmol/L | 3.83 | 4.74 | 0.89 | 2.19 |
| TC, mmol/L | 5.94 | 6.83 | 2.05 | 3.82 |
| non-fasting glucose, mmol/L | 5.78 | 6.21 | 7.85 | 5.15 |
| Statin | no | no | yes | yes |
| ARB | yes | no | yes | yes |
| ACE | no | no | no | no |
| β-blocker | no | no | yes | yes |
| CCB | yes | yes | yes | yes |
| Therapeutic anticoagulation | no | no | yes | no |
| Diuretics | yes | yes | yes | yes |
| ASA | no | no | no | yes |

**Supplementary Table 11: Primers used for gene expression analysis by quantitative real-time PCR (qPCR). Abbreviations:** ACTA2, alpha-smooth muscle actin; B2M, beta-2-microglobulin; COL1A1, collagen type I alpha 1 chain; COL1A1, collagen type I alpha 1 chain; COL3A1,collagen type III alpha 1 chain; E2F, E2F transcription factor 1; IL6, interleukin-6; NOS3, nitric oxide synthase 3 = endothelial nitric oxide synthase; RPL32, ribosomal protein L32.

| **Gene** | **Primers** | **Sequence, 5'-3'** |
|---|---|---|
| **NOX4** | Forward | CTTTTATCCAACAATCTCCTGGTTCTC |
| | Reverse | TAACCTCAACTGCAGCCTTATC |
| **RPL32** | Forward | CACCGTCCCTTCTCTCTTCCT |
| | Reverse | TCTTGGGCTTCACAAGGGGT |
| **B2M** | Forward | GATGAGTATGCCTGCCGTGT |
| | Reverse | CATGATGCTGCTTACATGTCTCG |
| **COL3A1** | Forward | CCTGAAGCTGATGGGGTCAA |
| | Reverse | TAGTCTCACAGCCTTGCGTG |
| **COL1A1** | Forward | AGACAGTGATTGAATACAAAACCA |
| | Reverse | GGAGTTTACAGGAAGCAGACA |
| **IL6** | Forward | CCTGACCCAACCACAAATGC |
| | Reverse | ATCTGAGGTGCCCATGCTAC |
| **NOS3** | Forward | GAACCTGTGTGACCCTCACC |
| | Reverse | TGGCTAGCTGGTAACTGTGC |

**Supplementary Table 12.** Concentrations of primary antibodies used for immunohistochemistry. Abbreviations: α-SMA, alpha-smooth muscle actin.

| **Protein** | **Order number and company** | **Used concentration** | **Duration of incubation** |
|---|---|---|---|
| α-SMA | A5228, Clone 1A4, Sigma Aldrich | 2 µg/mL | 1 h at RT |
| Cleaved caspase-3 | #9661, Cell Signaling | 0.59 µg/mL | 1 h at RT |
| CD31 | M0823, Clone JC70A, Agilent | 4.1 mg/L | 1 h at RT |
| CD68 | M0814, Clone KP1, Agilent | 4 µg/mL | overnight at 4°C |

**Supplementary Table 13.** Antibodies used for protein expression analysis by Western blot. Abbreviations: Erk, extracellular receptor kinase; MAPK, mitogen activated kinase; JNK, c-Jun N-terminal kinase

| **Protein** | **Company, Cat. #** | **Quantified size, kDa** |
|---|---|---|
| phospho-p38MAPK, Threonine$^{180}$/Tyrosine$^{182}$ | Cell Signaling, 9211 | 43 |
| Total p38MAPK | Cell Signaling, 9212 | 43 |
| Phospho-Smad3 Serine$^{423/425}$ | Cell Signaling, 9520 | 52 |
| Smad3 | Cell Signaling, 9523 | 52 |
| Phospho-SAPK/JNK, Threonine$^{183}$/Tyrosine$^{185}$ | Cell Signaling, 9255 | Double band at 46 and 54 |
| Total SAPK/JNK | Cell Signaling, 9252 | Double band at 46 and 54 |

#### References


1. Hofmann A, Muglich M, Wolk S, Khorzom Y, Sabarstinski P, Kopaliani I, et al. Induction of heme oxygenase-1 is linked to the severity of disease in human abdominal aortic aneurysm. J Am Heart Assoc. 2021;10(20):e022747.
2. Hofmann A, Khorzom Y, Klimova A, Wolk S, Busch A, Sabarstinski P, et al. Associations of Tissue and Soluble LOX-1 with Human Abdominal Aortic Aneurysm. J Am Heart Assoc. 2023;12(14):e027537.
3. Zheng GX, Terry JM, Belgrader P, Ryvkin P, Bent ZW, Wilson R, et al. Massively parallel digital transcriptional profiling of single cells. Nat Commun. 2017;8:14049.
4. Hao Y, Hao S, Andersen-Nissen E, Mauck WM, 3rd, Zheng S, Butler A, et al. Integrated analysis of multimodal single-cell data. Cell. 2021;184(13):3573-87 e29.
5. Kuleshov MV, Jones MR, Rouillard AD, Fernandez NF, Duan Q, Wang Z, et al. Enrichr: a comprehensive gene set enrichment analysis web server 2016 update. Nucleic Acids Res. 2016;44(W1):W90-7.
6. Badia IMP, Velez Santiago J, Braunger J, Geiss C, Dimitrov D, Muller-Dott S, et al. decoupleR: ensemble of computational methods to infer biological activities from omics data. Bioinform Adv. 2022;2(1):vbac016.
7. Temprano-Sagrera G, Peypoch O, Soto B, Dilme J, Calsina Juscafresa L, Davtian D, et al. Differential Expression Analyses on Human Aortic Tissue Reveal Novel Genes and Pathways Associated With Abdominal Aortic Aneurysm Onset and Progression. J Am Heart Assoc. 2024;13(24):e036082.
8. Dobin A, Davis CA, Schlesinger F, Drenkow J, Zaleski C, Jha S, et al. STAR: ultrafast universal RNA-seq aligner. Bioinformatics. 2013;29(1):15-21.
9. Li B, Dewey CN. RSEM: accurate transcript quantification from RNA-Seq data with or without a reference genome. BMC Bioinformatics. 2011;12:323.
10. Li Y, Ren P, Dawson A, Vasquez HG, Ageedi W, Zhang C, et al. Single-Cell Transcriptome Analysis Reveals Dynamic Cell Populations and Differential Gene Expression Patterns in Control and Aneurysmal Human Aortic Tissue. Circulation. 2020;142(14):1374-88.
11. Schröder K, Zhang M, Benkhoff S, Mieth A, Pliquett R, Kosowski J, et al. Nox4 is a protective reactive oxygen species generating vascular NADPH oxidase. Circulation Research. 2012;110(9):1217-25.
12. Busch A, Bleichert S, Ibrahim N, Wortmann M, Eckstein HH, Brostjan C, et al. Translating mouse models of abdominal aortic aneurysm to the translational needs of vascular surgery. JVS Vasc Sci. 2021;2:219-34.
13. Okuno K, Torimoto K, Cicalese SM, Hashimoto T, Sparks MA, Rizzo V, et al. Smooth muscle angiotensin II type 1A receptor is required for abdominal aortic aneurysm formation induced by angiotensin II plus beta-aminopropionitrile. J Mol Cell Cardiol. 2023;176:55-7.
